# From Chaos to Care: Personalized AI for Early Cardiac Arrhythmia Warning

**DOI:** 10.64898/2026.04.08.26350403

**Authors:** Suvankar Halder, Christopher M. Kim, Vipul Periwal

## Abstract

Cardiac arrhythmias are abnormal heart rhythms characterized by disordered electrical dynamics that impair cardiac function and pose a major global burden of morbidity and mortality. Early and accurate prediction of arrhythmic anomalies from physiological time series is crucial for effective intervention, yet remains challenging due to the nonlinear, nonstationary, and individualized nature of cardiac dynamics. Despite significant advances in machine learning-based arrhythmia detection, most existing methods operate as static classifiers on electrocardiographic signals and lack online prediction, patient-specific adaptation, and mechanistic interpretability.

From a dynamical-systems perspective, arrhythmias represent qualitative regime transitions, often preceded by subtle, temporally extended deviations that are difficult to detect in real time. Here we introduce CASCADE (Chaotic Attractor Sensitivity for Cardiac Anomaly Detection), an online and personalized anomaly forecasting framework built on a special type of reservoir computing called Dynamical Systems Machine Learning (DynML). DynML employs ensembles of continuous-time nonlinear dynamical systems as chaotic reservoirs to reconstruct and forecast short-term cardiac dynamics on a beat-to-beat basis, training only a linear readout. This design enables efficient online adaptation without retraining the underlying dynamical model. Rather than relying on static beat-level classification, CASCADE identifies arrhythmic events as failures of short-term predictability, manifested as statistically significant deviations between predicted and observed dynamics relative to subject-specific baselines.

Detection performance is governed by the intrinsic dynamical complexity of the reservoir, quantified by topological entropy. Reservoirs operating near critical entropy regimes optimally amplify subtle, temporally extended irregularities in heartbeat dynamics, rendering incipient arrhythmic signatures linearly separable at the readout level. Topological entropy thus serves both as a predictor of model performance and a principled control parameter for reservoir design. When evaluated on the MIT-BIH Arrhythmia dataset, CASCADE achieved consistently high F1 scores, precision, recall, and overall accuracy across diverse patient populations, demonstrating strong generalizability across clinical and real-world settings. By integrating chaotic reservoir computing, entropy-guided tuning, and online personalized forecasting, CASCADE reframes arrhythmia detection as a problem of dynamical regime transition rather than static classification. This perspective provides a scalable, interpretable, and computationally efficient framework for real-time cardiac monitoring and early-warning clinical decision support.

## 1 Introduction

Cardiac arrhythmias arise from complex spatiotemporal instabilities in the electrical activity of the heart and represent a major source of morbidity and mortality worldwide. From a dynamical-systems perspective, healthy cardiac rhythms correspond to stable or weakly perturbed oscillatory regimes, whereas arrhythmias reflect qualitative transitions to disordered, intermittently chaotic, or multistable dynamics driven by nonlinear interactions across molecular, cellular, and tissue scales [1–3]. Detecting and predicting such transitions from physiological time series remains a fundamental challenge, as arrhythmic events often emerge abruptly from subtle dynamical changes that precede overt pathological behavior. Critically, such transitions are rarely abrupt in state space; instead, they are often preceded by subtle dynamical deviations that remain difficult to detect in real time from physiological time series.

Traditional approaches to arrhythmia detection rely on handcrafted features extracted from electrocardiographic (ECG) signals, such as heart rate variability metrics, spectral measures, and morphological descriptors [4, 5]. While effective for retrospective classification, these methods struggle to capture the underlying nonlinear cardiac dynamics and typically lack predictive power for early anomaly detection. More recently, deep learning models—including convolutional and recurrent neural networks—have demonstrated strong performance on large annotated ECG datasets [6–8]. However, these approaches are fundamentally data-driven, require extensive labeled training data, and often fail to generalize across patients, recording conditions, or distinct dynamical regimes. Moreover, their internal representations are difficult to interpret in terms of cardiac physiology or underlying dynamical state transitions. Although such models achieve high accuracy in offline benchmarks, they are not explicitly designed for online prediction of future cardiac dynamics, nor do they provide mechanistic insight into how pathological rhythms emerge from patient-specific baseline dynamics. In particular, most existing approaches operate as static classifiers rather than predictive dynamical systems, limiting their ability to anticipate arrhythmic events before they manifest clinically [9–13].

From the standpoint of nonlinear dynamics, cardiac electrophysiology exhibits hallmark features of deterministic chaos, including sensitive dependence on initial conditions, multiscale temporal organization, and intermittent regime switching [14, 15]. These properties suggest that effective early-warning systems for arrhythmia should be framed not as pattern-recognition problems, but as problems of dynamical forecasting and regime identification. In this context, prediction of the near-future cardiac state becomes a prerequisite for meaningful anomaly detection. Reservoir computing (RC) provides such a framework by harnessing the transient dynamics of nonlinear systems to embed input signals into high-dimensional state spaces where complex temporal patterns become linearly separable [16, 17]. Unlike fully trained recurrent neural networks, reservoir computing trains only a linear readout, enabling efficient learning while preserving the intrinsic dynamics of the reservoir.

Recent work has shown that physical and dynamical reservoirs—including chaotic oscillators, delay systems, and continuous-time nonlinear flows—can perform computation by leveraging their natural evolution [18, 19]. These systems are particularly well suited for processing physiological signals, which themselves arise from nonlinear biological dynamics. Nevertheless, a principled understanding of which reservoir dynamics are most effective for anomaly detection remains incomplete. In particular, the role of intrinsic dynamical complexity—quantified by measures such as Lyapunov exponents or topological entropy—has not been systematically linked to predictive performance in physiological time series.

In our previous work [20], we introduced Dynamical Systems Machine Learning (DynML), a reservoir computing framework that integrates continuous-time nonlinear dynamical systems with data-driven readouts to model complex biological dynamics. DynML exploits the intrinsic transient evolution of chaotic reservoirs to encode temporal structure while training only a linear readout, enabling computational efficiency and mechanistic interpretability. Importantly, we demonstrated that the predictive performance of DynML is strongly correlated with the topological entropy of the underlying reservoir dynamics, establishing a quantitative link between dynamical complexity and model expressivity.

In the present work, we extend DynML to online, personalized prediction and anomaly detection in cardiac electrophysiology through a new architecture termed CASCADE (Chaotic Attractor Sensitivity for Cardiac Anomaly Detection). Rather than directly classifying arrhythmic beats, CASCADE continuously forecasts the short-term evolution of cardiac time series on a beat-to-beat basis using a nonlinear dynamical reservoir with a linear readout, and identifies arrhythmic anomalies as statistically significant deviations between predicted and observed dynamics relative to patient-specific baselines. As summarized schematically in Fig. 1, CASCADE analyzes ECG signals through a sequence of interpretable steps designed for continuous, real-time monitoring. First, short segments of each heartbeat are represented using sliding windows and compressed into a low-dimensional, patient-specific state space that captures the dominant patterns of normal cardiac activity. Next, a dynamical learning module (DynML) is trained to predict the immediate future signal within each segment based on its recent history, effectively learning the underlying temporal structure of a patient’s baseline rhythm. The discrepancy between predicted and observed signals (prediction error) is then continuously evaluated, and its statistical distribution under normal conditions is estimated from validation data. During deployment, the model monitors deviations from this learned error distribution: when prediction errors become systematically unlikely under the normal regime, the system flags a transition away from baseline dynamics. By framing arrhythmia detection as the identification of dynamical regime transitions rather than static classification, this approach naturally accommodates inter-patient variability and avoids reliance on rigid class boundaries. The resulting framework is physically grounded, interpretable, and well-suited for robust, online anomaly detection in complex physiological time series.

**Figure 1:**
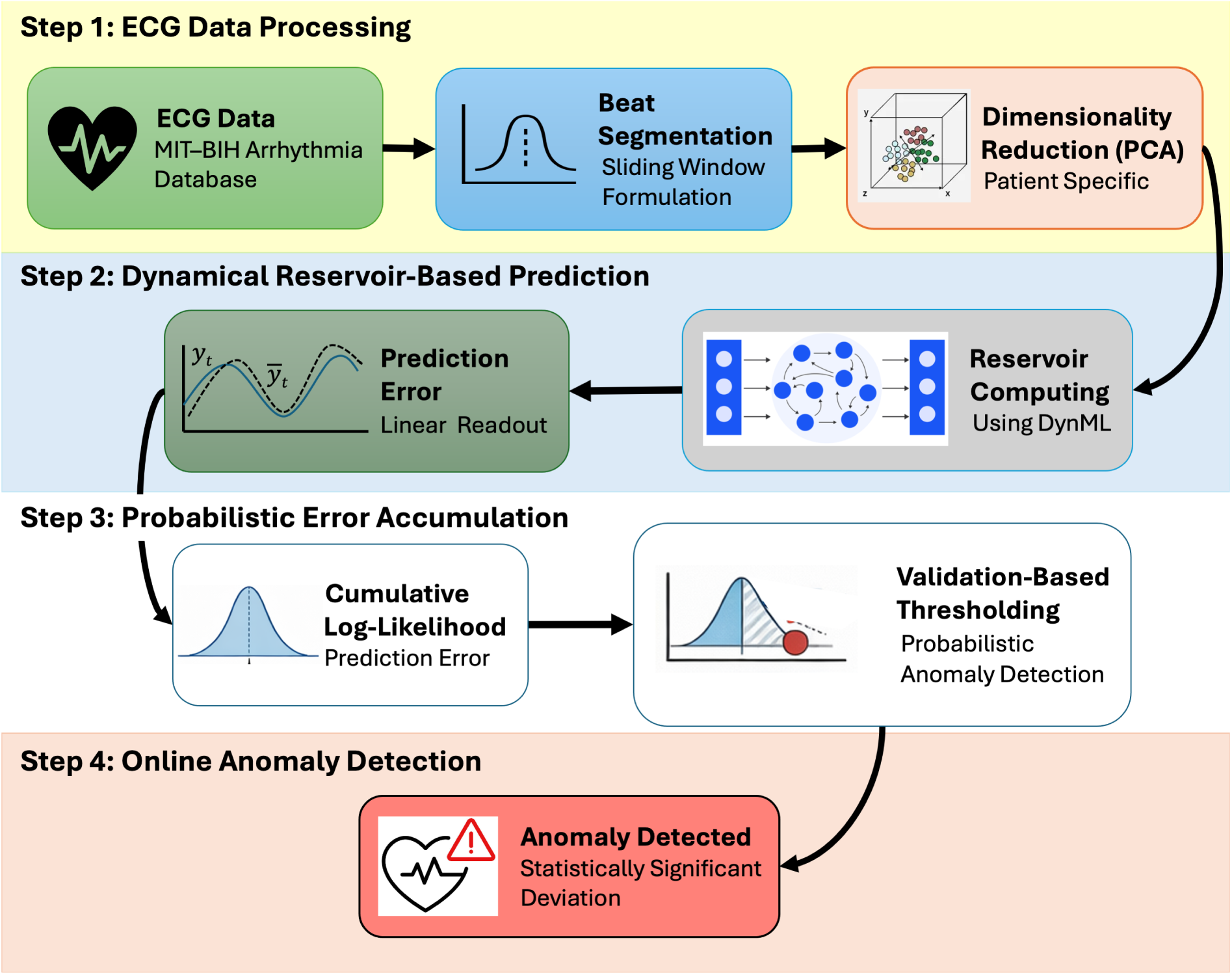
Schematic overview of the CASCADE framework. The workflow consists of four main stages: **Step 1: ECG data processing**, where ECG signals are segmented into beat-level sliding windows and projected into a patient-specific low-dimensional PCA space; **Step 2: Dynamical reservoir-based prediction**, where the reduced signals drive a nonlinear reservoir operating near critical regimes of topological entropy and a linear readout performs one-step-ahead forecasting; **Step 3: Probabilistic error accumulation**, where prediction errors are modeled using validation-derived Gaussian reference distributions and aggregated through cumulative log-likelihoods; and **Step 4: Online anomaly detection**, where cumulative deviation statistics are compared against patient-specific thresholds to identify arrhythmic events in real time.

A key contribution of this study is the explicit role of reservoir dynamical complexity in enabling early and reliable anomaly detection. We show that nonlinear reservoirs operating near critical regimes of topological entropy optimally amplify subtle irregularities in cardiac dynamics, rendering incipient arrhythmic signatures linearly separable at the readout level. Topological entropy thus emerges not only as a predictor of DynML performance [20], but also as a principled control parameter for designing reservoir systems optimized for online detection of dynamical regime shifts. Because DynML trains only the readout layer, such entropy-tuned systems can efficiently adapt to individual patients while preserving sensitivity to clinically relevant deviations.

## 2 Methods

### 2.1 Data Acquisition

ECG data used in this study were obtained from the MIT–BIH Arrhythmia Database [5], a widely adopted benchmark dataset for the evaluation of cardiac arrhythmia detection algorithms.

The database includes long-term ECG recordings from a diverse cohort of subjects, with expert annotations identifying normal and arrhythmic beats. Signals are sampled at 360 Hz and include two ECG leads per subject, providing high temporal resolution for beat-to-beat analysis. The MIT–BIH database has been used in approximately 61% of deep learning–based arrhythmia studies [21], making it a standard reference for reproducible and comparable evaluations.

### 2.2 ECG Data and Beat Segmentation

Lead I signals were used for all analyses. Expert annotations were used to identify R-peak locations and beat types. Normal beats (N, L, R, e, j, .) and arrhythmic beats (A, a, J, S, V, E, F) were extracted following standard conventions [22]. Each heartbeat was represented as a fixed-length segment of 180 samples (corresponding to 500 ms at a sampling rate of 360 Hz) preceding the R-peak, as illustrated in Fig. 2, where the shaded region shows the selected beat segment preceding the R-peak. This window length was selected based on an empirical analysis of inter-beat interval distributions in the selected patient cohort (see Patient Selection), which showed that the majority of normal-to-normal, normal-to-arrhythmic, and arrhythmic-to-arrhythmic transitions exceeded this duration (Fig. 3). Choosing 180 samples ensures that each segment captures the complete preceding cardiac cycle while minimizing overlap with earlier beats. Beats with insufficient preceding samples were excluded. For each selected patient, normal beats were randomly partitioned into training, validation, and test subsets, while arrhythmic beats were reserved exclusively for testing. This strategy ensures that model training is based solely on patient-specific normal dynamics, enabling robust online anomaly detection.

**Figure 2:**
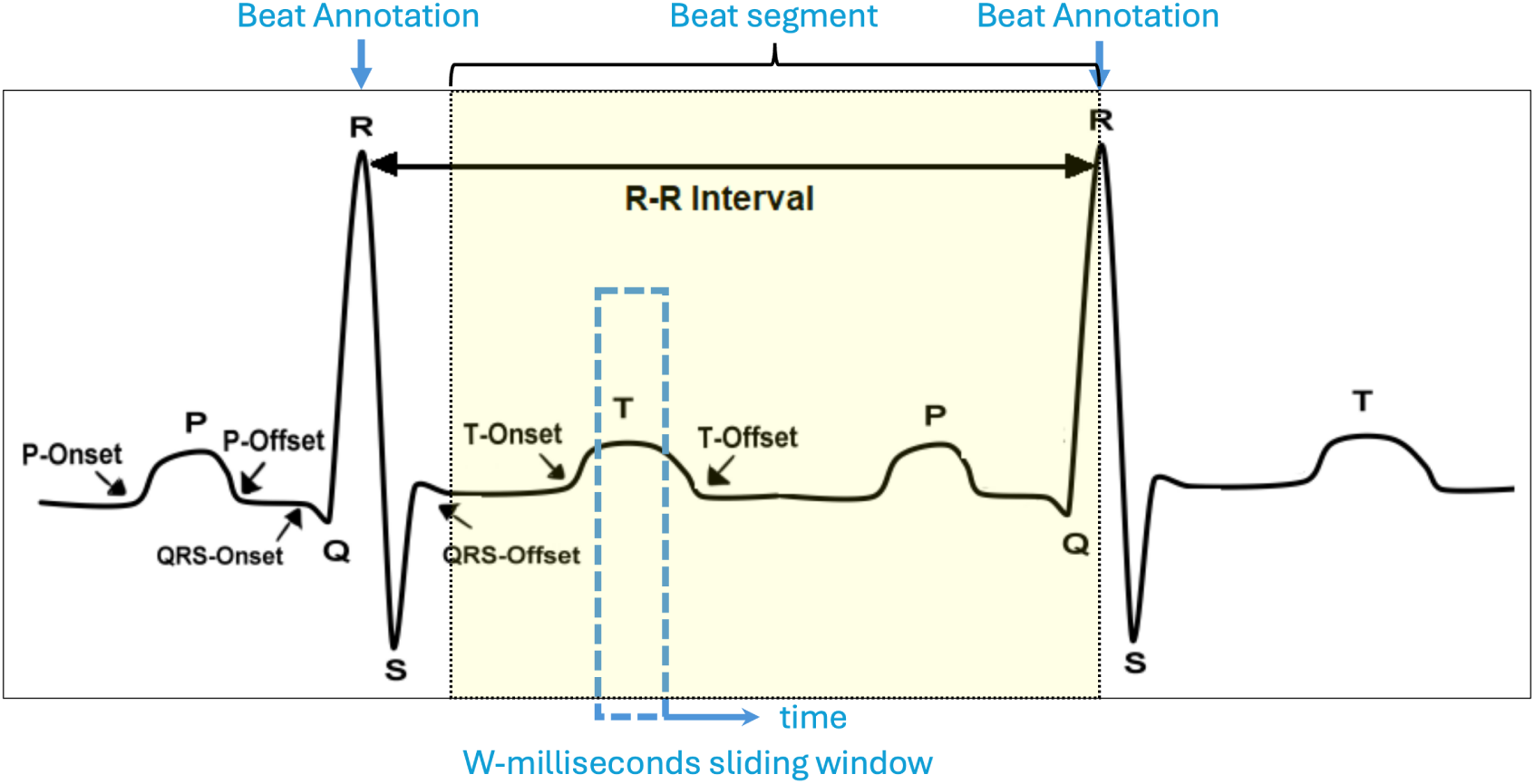
Beat segment definition and sliding window representation used in this study. The shaded region represents the selected ECG segment corresponding to a single beat, defined as a fixed-length window preceding the subsequent R-peak. Within this segment, a sliding window of length *W* is used to construct input sequences for next-sample prediction, as described in the sliding window formulation. Each window captures the preceding cardiac dynamics and is associated with the subsequent sample, while the segment terminates at an annotated beat (normal or arrhythmic), which serves as the prediction target.

**Figure 3:**
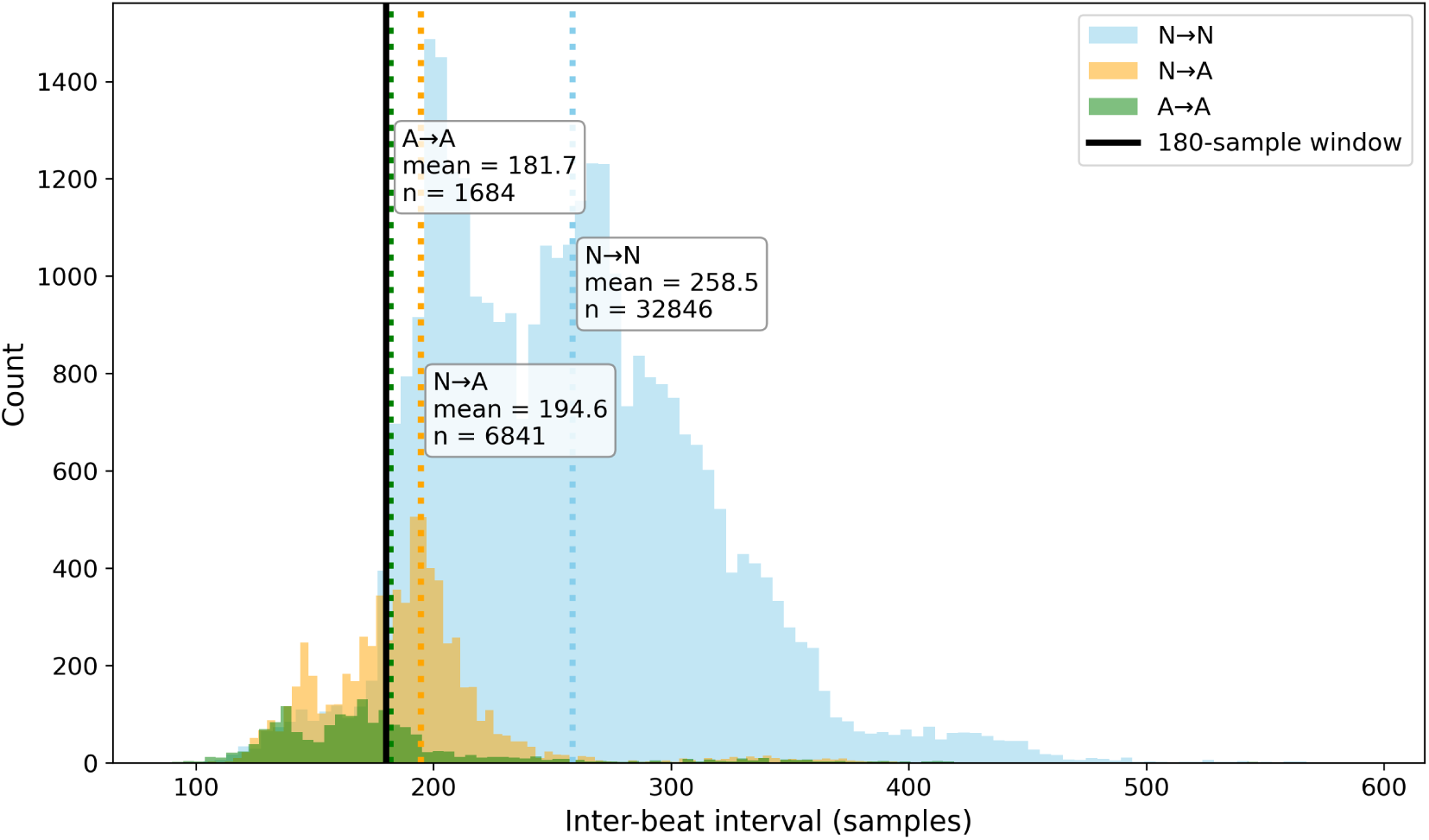
Distribution of inter-beat intervals (in samples) for selected transitions: normal-to-normal (*N* → *N* ), normal-to-arrhythmic (*N* → *A*), and arrhythmic-to-arrhythmic (*A* → *A*). Vertical dotted lines indicate the mean interval for each transition, and the solid black line shows the 180-sample window used for beat segmentation. This analysis supports the choice of 180 samples as a representative segment length that captures the majority of preceding cardiac cycles.

### 2.3 Patient Selection

Patients were selected from the dataset based on the availability of sufficient normal and arrhythmic beats to support reliable modeling and evaluation. Specifically, inclusion required at least 700 normal beats and 100 arrhythmic beats per patient, ensuring adequate representation of both baseline and pathological cardiac dynamics. For each selected patient, 500 normal beats were used for training and an additional 100 normal beats for validation, yielding a total of 600 normal beats for model fitting and parameter tuning. The test set comprised 100 previously unseen normal beats and 100 arrhythmic beats, enabling rigorous assessment of CASCADE’s online predictive and anomaly detection performance under both physiological and pathological conditions. Applying these criteria resulted in a cohort of 19 patients. Figure 4 shows the beat distributions for all 19 selected patients, highlighting inter-patient variability and confirming that all inclusion criteria were met. This patient selection and data partitioning strategy ensures a consistent, balanced, and clinically relevant evaluation of CASCADE across diverse cardiac profiles.

**Figure 4:**
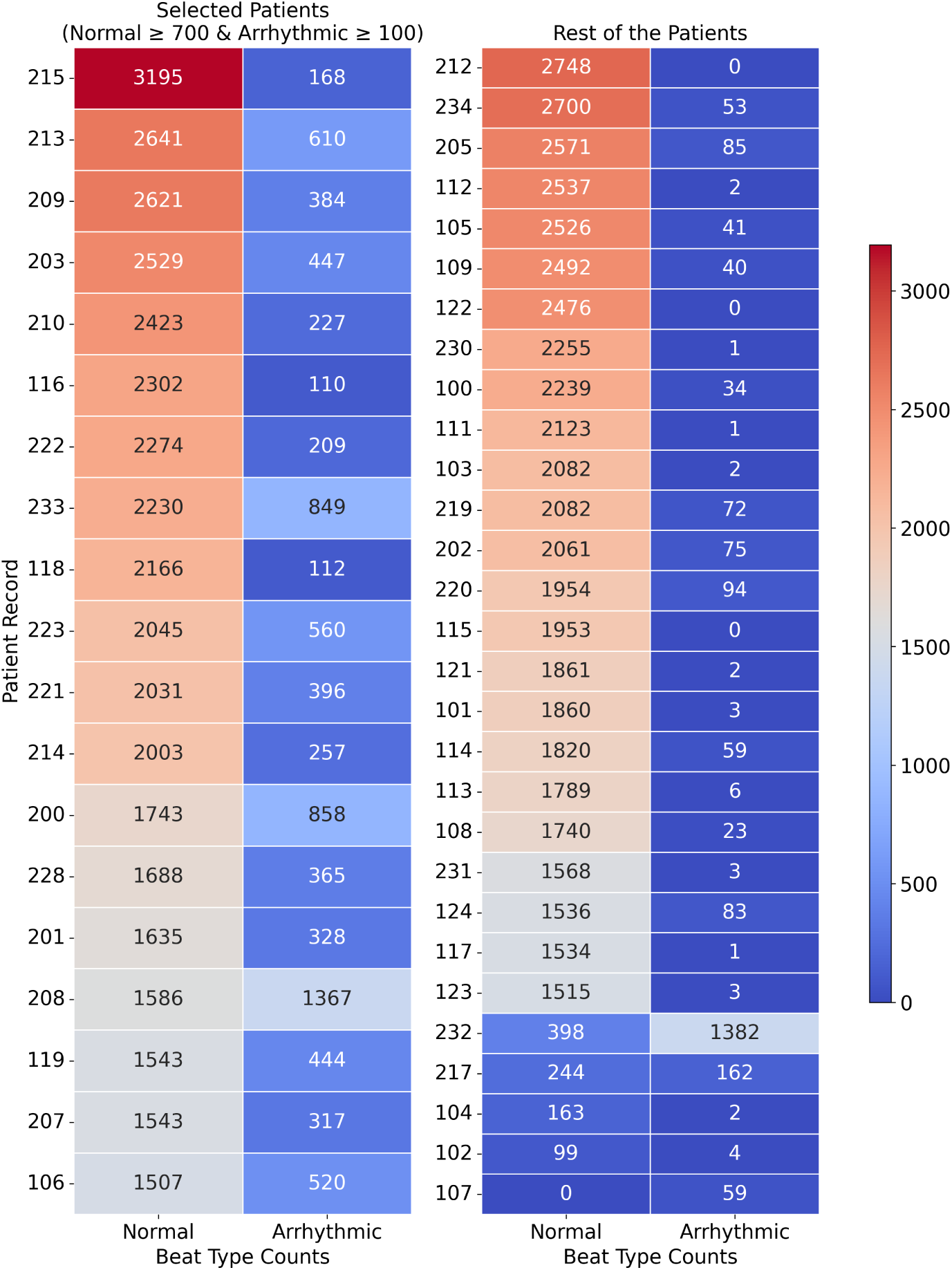
Patient selection based on beat counts for CASCADE. Patient records from the MIT-BIH dataset, showing inclusion based on sufficient numbers of normal and arrhythmic beats. The left panel displays selected patients with at least 700 normal beats and 100 arrhythmic beats, suitable for training, validation, and testing of CASCADE. The right panel shows patients excluded due to insufficient beat counts. Color intensity indicates the number of beats for each type, with the shared colorbar providing a consistent scale. This selection ensures adequate physiological data for reliable online anomaly detection and highlights variability in beat distributions across individuals.

### 2.4 CASCADE Framework

The CASCADE framework implements personalized, online arrhythmia forecasting using DynML. For each patient, a personalized baseline of normal cardiac dynamics is first established from normal beats. Individual heartbeat segments are then embedded into an ensemble of continuous-time nonlinear dynamical systems (Rössler units) operating near critical dynamical regimes, forming a patient-specific reservoir representation. A linear readout layer is trained to perform short-term, beat-to-beat forecasting of cardiac dynamics based on this embedding. During online operation, arrhythmic anomalies are detected by identifying statistically significant deviations between predicted and observed dynamics relative to subject-specific thresholds derived from normal behavior. The overall CASCADE pipeline is summarized schematically in Fig. 1, which illustrates the progression from beat segmentation and reservoir embedding to short-term prediction, probabilistic error modeling, and online anomaly detection. The following steps summarize the methodology:

### Sliding Window Formulation

Each heartbeat segment is divided into overlapping windows of fixed length *W* samples, as illustrated in Fig. 2, which depicts the sliding-window construction over a single beat segment. For each window, the goal is to predict the next ECG sample immediately following the window. Formally, for a given window of samples

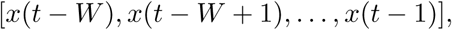

the model predicts the next sample *x*(*t*). Sliding this window along the segment generates multiple input-output pairs

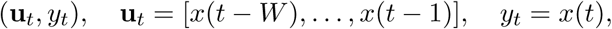

which are used for training and testing.

This formulation allows the model to perform online, beat-to-beat prediction and facilitates real-time detection of deviations from normal cardiac dynamics.

### Dimensionality Reduction

To reduce redundancy in the input windows and stabilize reservoir dynamics, principal component analysis (PCA) was applied as a preprocessing step. PCA was fit exclusively on the training data for each input window length and subsequently used to project the test windows, ensuring that no information from anomalous beats influenced the learned embedding.

For the selected cohort of patients, PCA was evaluated across multiple input window lengths (*W* = 10–50), with each setting producing well-sampled embeddings (up to ∼ 8.5 × 10^4^ training windows per patient). Across all patients and window lengths, three-dimensional PCA consistently captured the dominant variance structure, explaining over 92% of the total variance in the training data. In contrast, two-dimensional projections failed to capture sufficient variance for all patients, and four-dimensional PCA provided only marginal gains, reaching a maximum of ∼ 94% variance explained for some patients at select window lengths (Figure 5).

**Figure 5:**
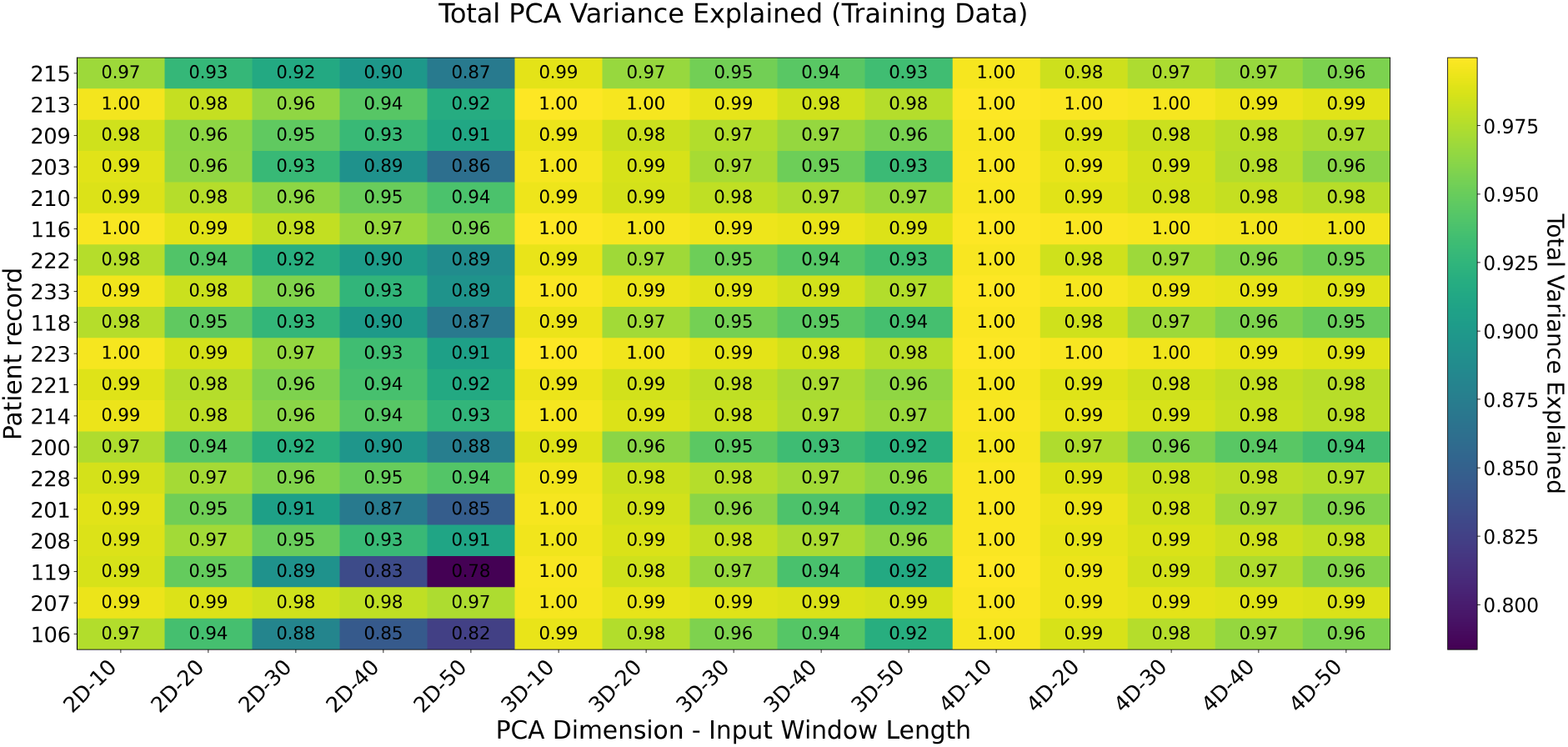
Total variance explained by PCA across patients and input window lengths (training data). Heatmap showing the proportion of variance captured by PCA for the selected patients. Columns represent all combinations of PCA dimensions (2D, 3D, 4D) and input window lengths (*W* = 10–50 samples), labeled as “PCA Dimension - Window Length” (e.g., 2D-10, 3D-20). Rows correspond to patients. Each cell value indicates the total variance explained by the given number of principal components on the training data. A single colorbar (right) reflects the magnitude of variance explained. Values are also annotated within each cell for clarity.

These results indicate that three principal components are sufficient to preserve the intrinsic structure of normal cardiac dynamics while maintaining discriminative capacity for arrhythmic deviations. For example, in Patient 106, the first three principal components of the training data captured over 92% of the variance even in the input window length with the least explained variance, and the corresponding 3D embeddings of the test windows revealed a clear separation between normal and arrhythmic beats: normal beats formed a compact manifold, while arrhythmic beats deviated distinctly along the principal axes (Figure 6). In contrast, 2D projections captured only about 82% of the variance in the worst-case window, resulting in substantial class overlap, while adding a fourth component increased the explained variance to approximately 96% for the least-explained window but did not meaningfully improve separability. These patterns were consistent across all selected patients and all tested input window lengths (Figure 5). Consequently, all input windows were projected into this three-dimensional latent space prior to reservoir embedding, achieving an optimal balance between dimensional efficiency, noise suppression, and variance retention.

**Figure 6:**
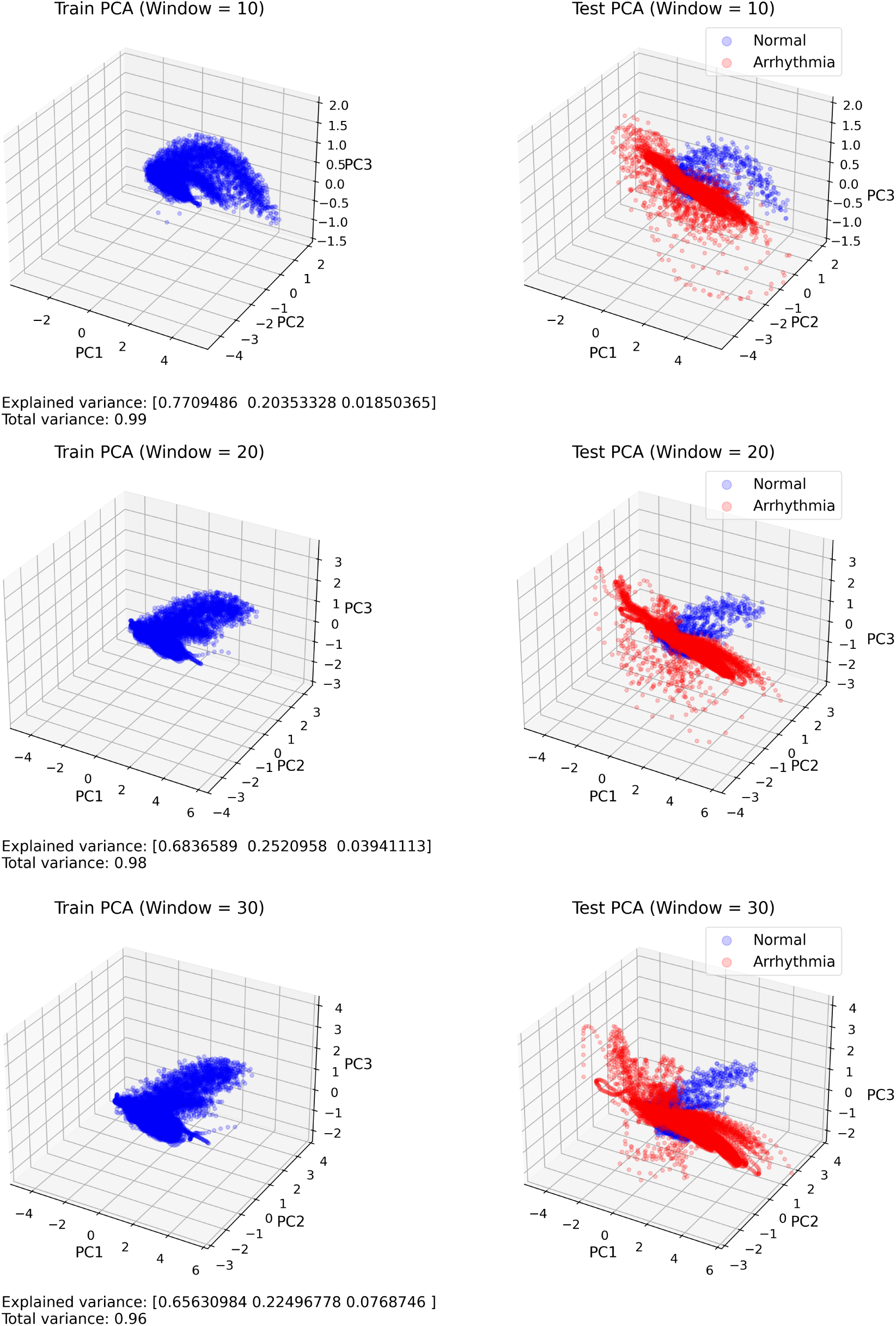
PCA-based separability of normal and arrhythmic ECG windows (Continued figure: Part 1). Three-dimensional PCA projections of beat-level ECG windows for a representative patient (Patient 106), shown across multiple input window lengths (*W* = 10–30). PCA was trained exclusively on normal beats, and test data—including both normal (blue) and arrhythmic (red) beats—were projected using the same transformation. Normal beats form a compact manifold, while arrhythmic beats deviate clearly along the first three principal components. The first three components capture the dominant variance in the training set (over 92%) and provide improved class separability in the test set.

**Figure 6:**
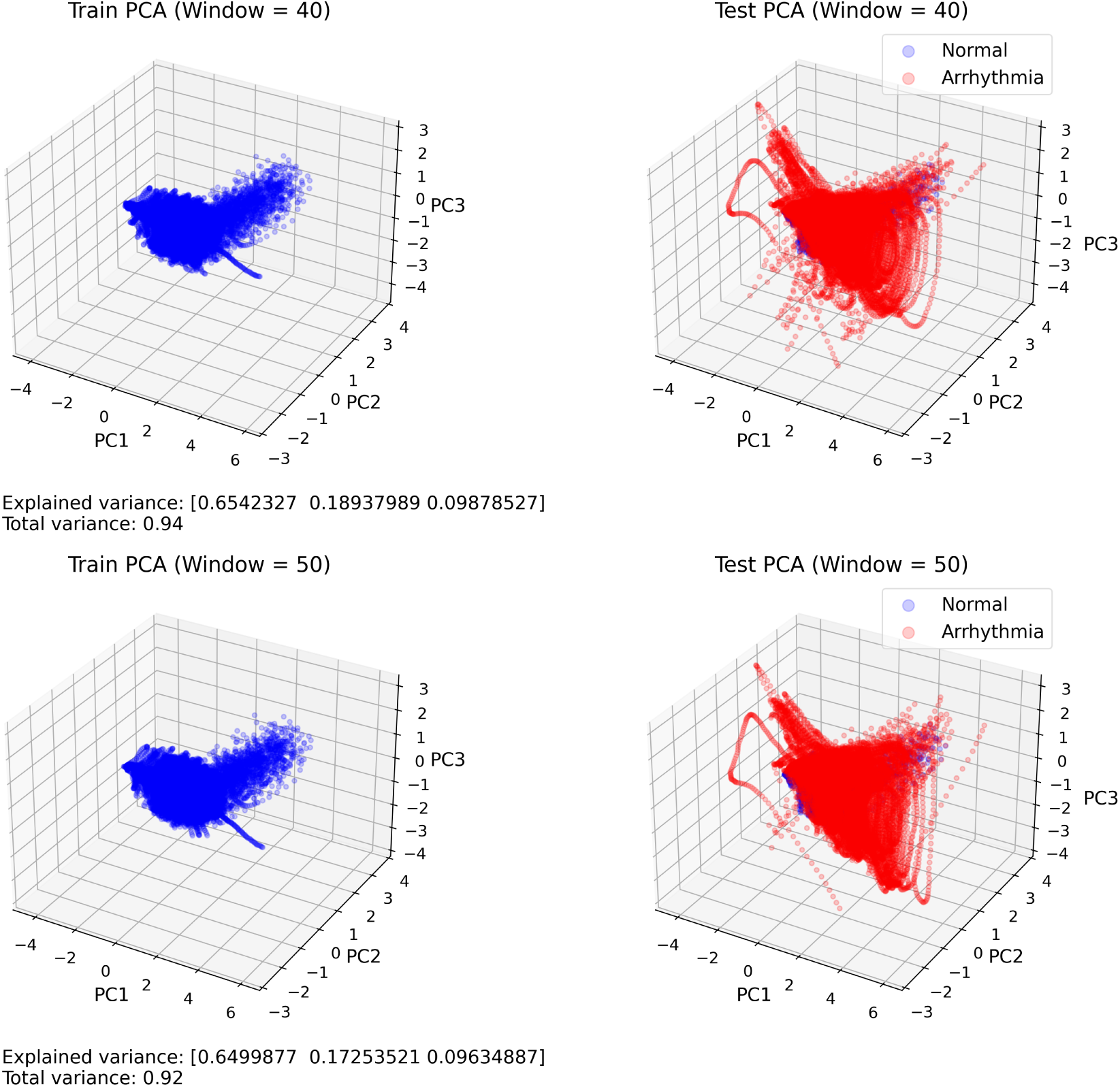
PCA-based separability of normal and arrhythmic ECG windows (Continued figure: Part 2(last)). Three-dimensional PCA projections of beat-level ECG windows for a representative patient (Patient 106), shown across multiple input window lengths (*W* = 40–50). PCA was trained exclusively on normal beats, and test data—including both normal (blue) and arrhythmic (red) beats—were projected using the same transformation. Normal beats form a compact manifold, while arrhythmic beats deviate clearly along the first three principal components. The first three components capture the dominant variance in the training set (over 92%) and provide improved class separability in the test set.

### Reservoir Dynamics

The core of the CASCADE framework consists of an ensemble of heterogeneous continuous-time nonlinear dynamical systems, forming the dynamical reservoir (for details see DynML paper [20]). Each reservoir unit is implemented as a Rössler system, with dynamics governed by:

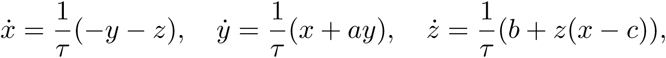

where (*a, b, c, τ* ) are parameters sampled from distributions spanning stable to chaotic regimes. The time constant *τ* scales the evolution of the reservoir states.

Input vectors extracted from each sliding ECG window are mapped into the reservoir through a fixed random linear projection, which initializes the reservoir state for that input. Specifically, each window is injected as a single input instance rather than as a continuously driven time series. The reservoir dynamics are then evolved autonomously from this initialized state using numerical integration of the governing nonlinear system over a fixed time horizon. The terminal state of this evolution is taken as the reservoir representation of the input, yielding a high-dimensional embedding that reflects the intrinsic dynamical response of the system to the input perturbation. By repeating this process across successive overlapping windows, the reservoir effectively captures temporal dependencies in the signal, providing a dynamical memory that supports short-horizon prediction of ECG waveform evolution.

### Linear Readout and Prediction

A linear readout layer is trained on normal beats to map reservoir states to predicted ECG values for the next time step. During inference, the trained model generates one-step-ahead predictions in an online fashion.

### Statistical Characterization of Prediction Errors

For each time step *t* within a heartbeat, the model generates a one-step-ahead prediction *ŷ_t_*, which is compared against the observed signal value *y_t_* to yield a prediction error *ɛ_t_* = *y_t_* − *ŷ_t_*. We analyzed these prediction errors using normal beats from a held-out validation set to characterize their statistical structure under normal cardiac dynamics. Errors were standardized using time-dependent variance estimates computed from the validation data to enable aggregation across time points and beats.

As shown in Fig. 7a, the histogram of standardized prediction errors closely follows a Gaussian distribution fitted using the empirical mean and standard deviation, indicating that the residuals are approximately symmetric and bell-shaped. This observation is further supported by the quantile–quantile (Q–Q) plot in Fig. 7b, which compares standardized residuals to a standard normal distribution (*µ* = 0, *σ* = 1) and demonstrates near-linear alignment, indicating minimal skewness and absence of heavy-tailed behavior.

**Figure 7:**
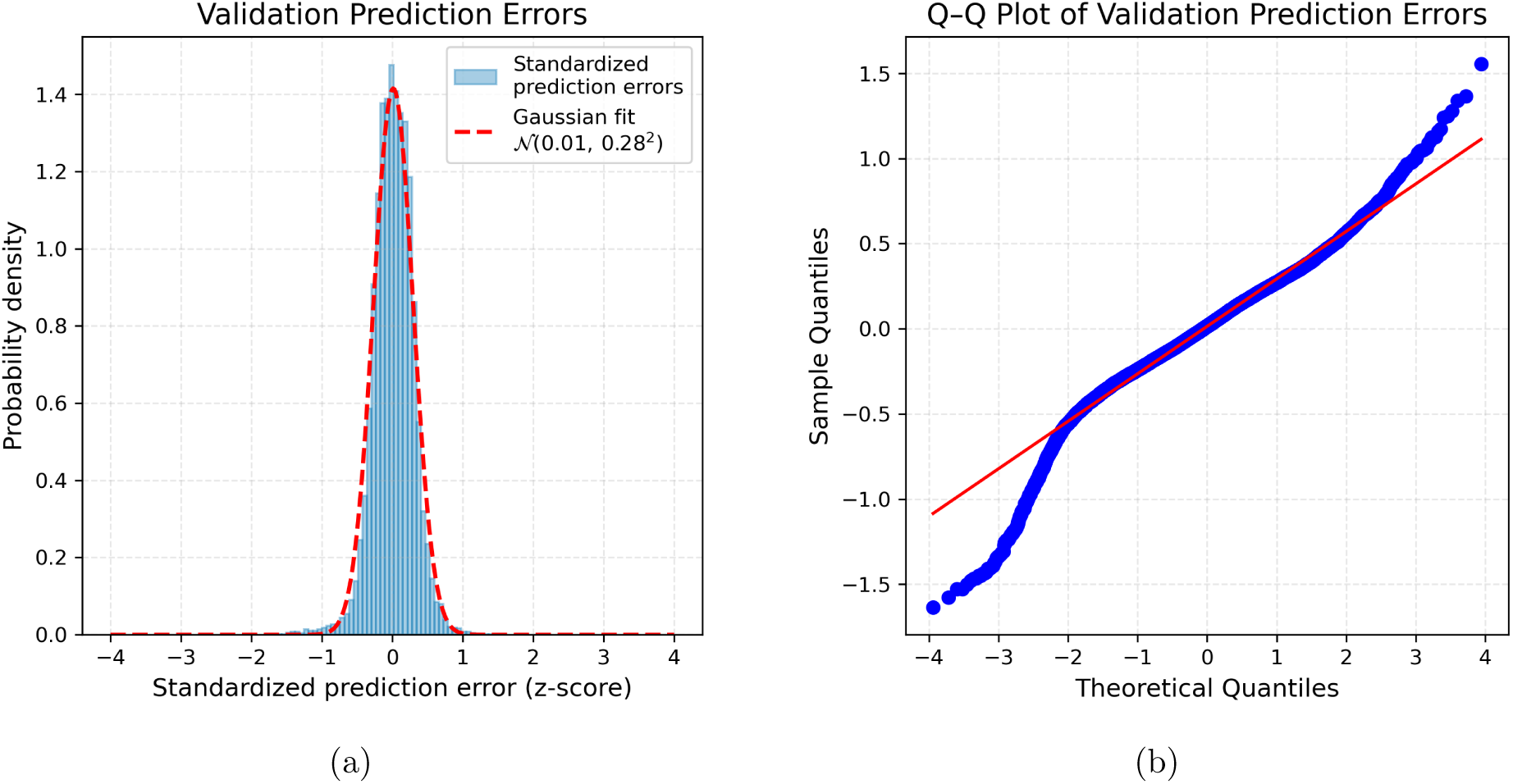
Statistical structure of prediction errors under normal cardiac dynamics. (**a**) Histogram of standardized prediction errors from normal beats in the validation set. The dashed red curve shows a Gaussian probability density function fitted to the empirical mean and standard deviation of the errors, highlighting the approximate bell-shaped distribution of the residuals. (**b**) Quantile–quantile (Q–Q) plot comparing standardized validation-set residuals with a standard normal distribution (*µ* = 0, *σ* = 1). The near-linear alignment indicates that the standardized errors closely follow Gaussian statistics, supporting the use of a Gaussian likelihood for anomaly detection.

Together, these empirical observations indicate that prediction errors under normal cardiac dynamics are well-approximated by a Gaussian distribution. This data-driven characterization motivates the use of a Gaussian likelihood, which forms the statistical foundation of the subsequent likelihood-based anomaly detection framework. All likelihood parameters and decision thresholds are derived exclusively from the validation set and fixed prior to testing.

### Validation-Based Thresholding

Motivated by the empirical Gaussian structure of validation-set prediction errors, we construct patient-specific reference distributions from normal beats to detect arrhythmic anomalies (Figure 8). For each time step *t* within a beat, the predicted value *ŷ_t_* is compared against the true observation *y_t_*, yielding a prediction error *ɛ_t_* = *y_t_* − *ŷ_t_*. Based on the validation analysis, these errors are modeled as Gaussian,

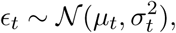

where *µ_t_*and *σ_t_* denote the mean and standard deviation of prediction errors at time *t*, estimated exclusively from validation data.

**Figure 8:**
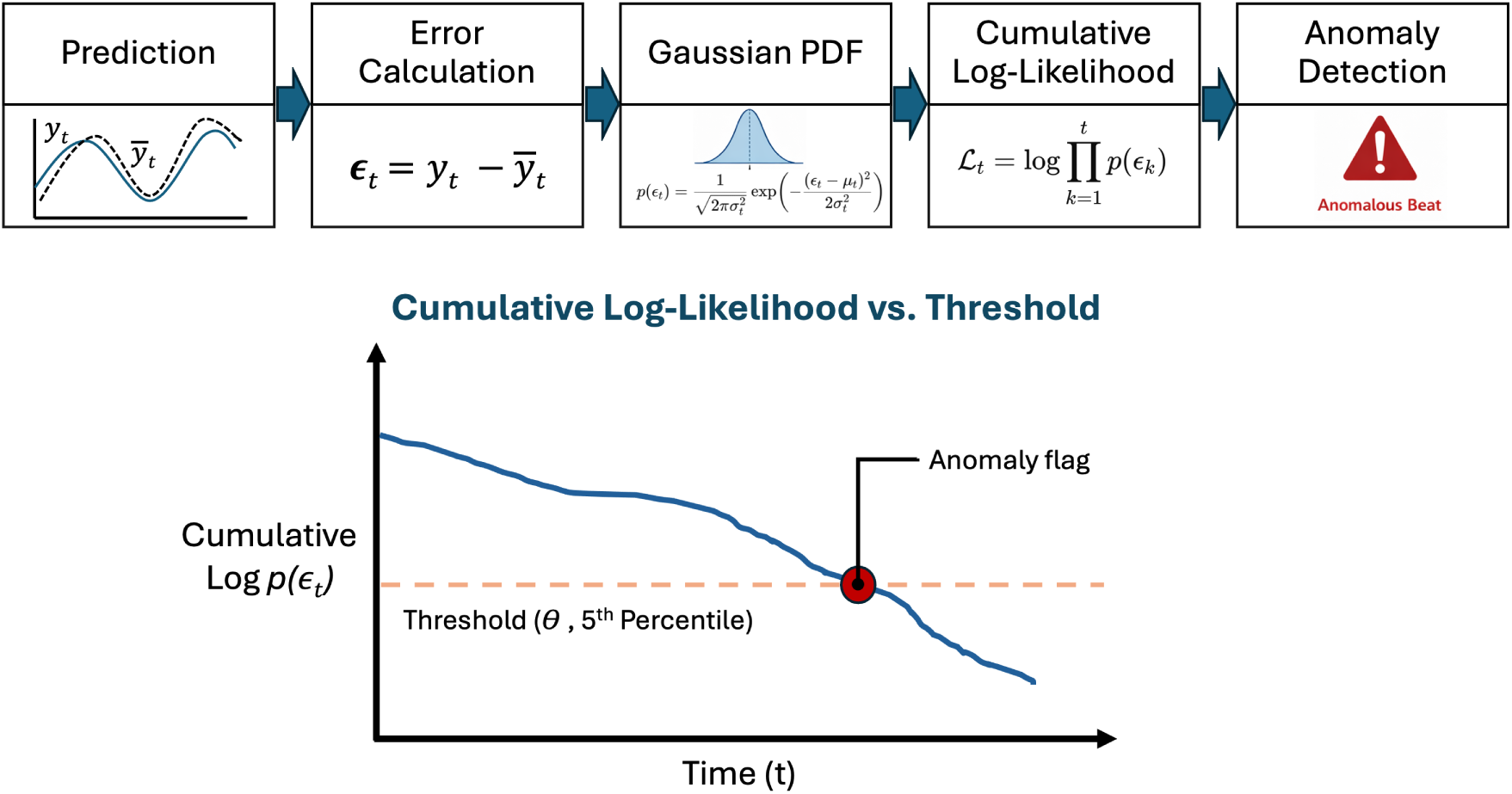
Validation-based probabilistic anomaly detection framework. Prediction errors *ɛ_t_* = *y_t_* − *ŷ_t_* obtained from normal validation beats are used to estimate time-dependent Gaussian reference distributions with mean *µ_t_* and standard deviation *σ_t_*. During testing, the probability density of each observed error is evaluated under the corresponding Gaussian model, and log-likelihoods are accumulated over time to form a cumulative log-probability score. Patient-specific anomaly thresholds *θ_t_* are defined as a lower percentile (e.g., 5th percentile) of cumulative log-likelihoods computed on validation data. A beat is flagged as arrhythmic when its cumulative log-likelihood falls below the threshold, indicating a statistically significant deviation from normal cardiac dynamics.

The probability density function (PDF) of the Gaussian distribution provides a measure of how likely a given prediction error is:

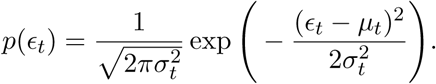

Intuitively, smaller likelihoods indicate deviations from the expected normal behavior. To aggregate evidence over multiple time points within a beat, we compute the cumulative log-likelihood:

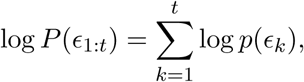

which quantifies the joint probability of the observed sequence of errors under the normal-beat model. Low cumulative probability indicates that the sequence is unlikely under normal conditions, suggesting an arrhythmic deviation.

A patient-specific threshold *θ*(*t*) is derived from the distribution of cumulative log-likelihoods on the validation set, typically using a lower percentile (e.g., 5th percentile) to control false positive rates:

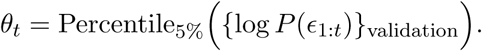

During online testing, each new beat is projected into this framework, and the cumulative log-likelihood is continuously monitored. A beat is flagged as anomalous if

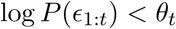

for any *t*, indicating that the observed deviations are statistically unlikely under the learned normal dynamics.

This probabilistic approach leverages the assumption that normal cardiac dynamics are well-characterized by the Gaussian reference distributions derived from validation data. By accumulating evidence over time, the method captures both transient and sustained deviations, enabling robust detection of arrhythmic beats while minimizing false positives caused by natural physiological variability. The framework is flexible, patient-specific, and naturally integrates the time-resolved nature of prediction errors for sequential anomaly detection. A schematic overview of the validation-based probability estimation, cumulative evidence accumulation, and threshold-based anomaly decision is shown in Figure 8.

### Online Anomaly Detection

During testing, cumulative log-likelihoods were updated in real time. A beat was flagged as anomalous if its cumulative log-likelihood dropped below the validation-derived threshold. Performance was quantified over time using accuracy, precision, recall, and F1 score.

All components of the framework, including the reservoir dynamics, readout mapping, and anomaly thresholds, are patient-specific. This ensures that natural variations in cardiac dynamics do not trigger false positives, providing robust and individualized arrhythmia detection.

## 3 Results

### 3.1 Online Prediction Performance of CASCADE

We evaluated the online predictive performance of the CASCADE framework for next-point ECG prediction across multiple input window lengths for all selected patients. The F1 score, which quantifies the accuracy of online arrhythmic event detection, was computed at each predicted time step following the input window. As shown for a representative subject (Patient 106) in Fig. 9, the F1 score across all input windows begins near 0.8 at the earliest prediction points and increases progressively as more points are incorporated into the probability estimates used for classification. Notably, a pronounced improvement in F1 score is consistently observed after approximately 200 ms, beyond which performance rapidly improves and approaches saturation. This temporal evolution follows a characteristic sigmoid-like trajectory, indicating that the model accumulates sufficient predictive evidence beyond this timescale to reliably confirm arrhythmic dynamics.

**Figure 9:**
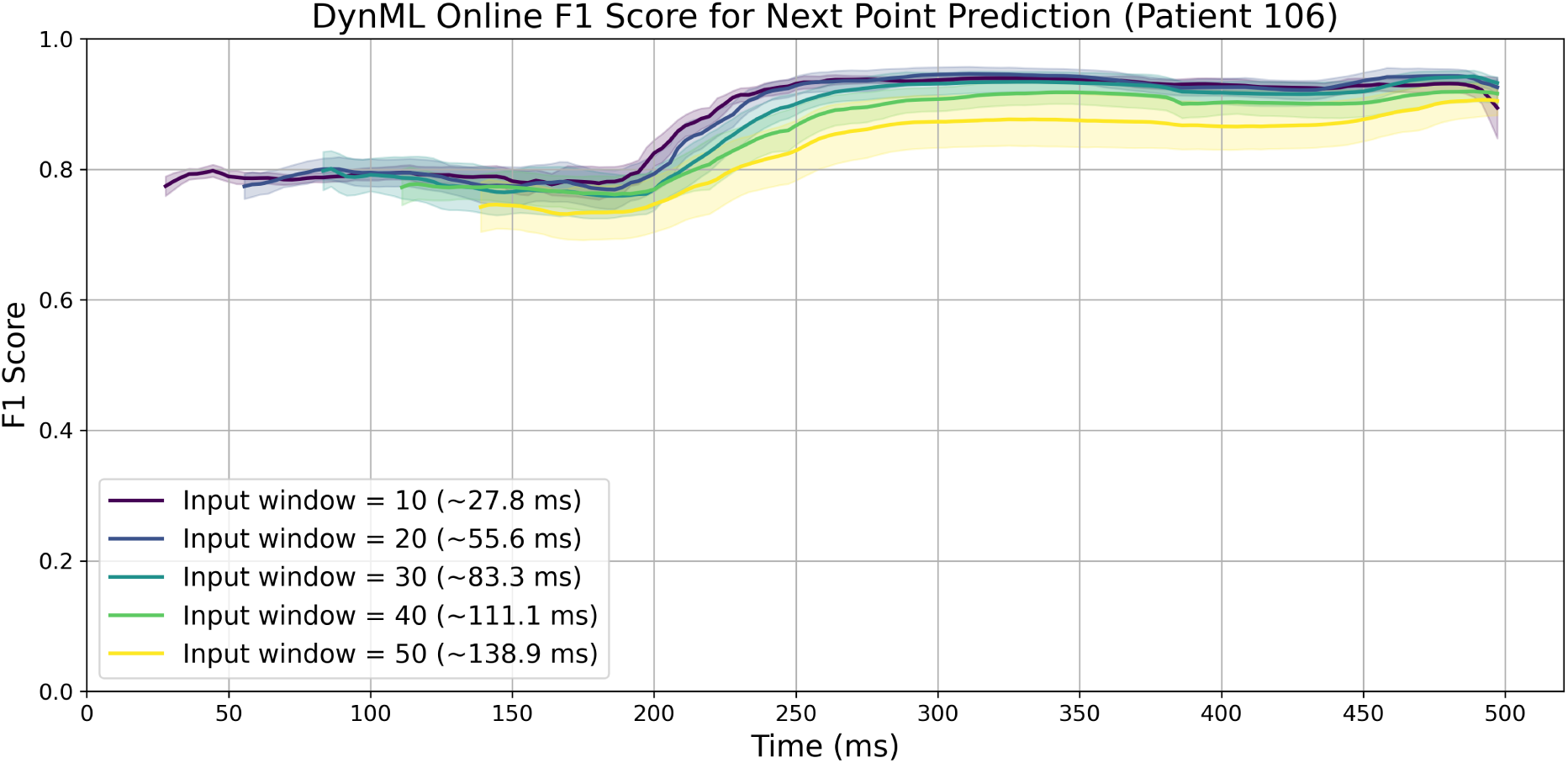
CASCADE online F1 score for next-point ECG prediction in Patient 106. Mean F1 score trajectories are shown for different input window lengths (10–50 samples) as a function of predicted time (ms). Results are averaged over ten independent random seeds for the DynML model, with shaded regions indicating ± one standard deviation. Across all window sizes, the F1 score begins near 0.8 and increases with prediction time, exhibiting a sharp rise after approximately 200 ms before saturating near 1.0. Longer input windows start with slightly lower initial F1 scores but achieve comparable final performance, illustrating the trade-off between early detection and temporal integration in the CASCADE framework.

Longer input windows tend to exhibit lower initial F1 scores but converge toward saturation near the theoretical maximum of 1 as more prediction steps are considered. This behavior arises because, although longer input windows provide richer temporal context for standard time-series prediction, our framework relies on prediction error for anomaly detection rather than prediction accuracy alone. This behavior suggests a trade-off between early responsiveness and long-term confidence: shorter input windows enhance sensitivity to small deviations due to the chaotic reservoir dynamics, leading to larger prediction errors and earlier threshold crossing for anomaly detection. In contrast, longer input windows stabilize the reservoir dynamics, improving time-series prediction accuracy while reducing early prediction error, thereby delaying anomaly detection.

Importantly, similar trends in the temporal evolution and saturation of online F1 scores were consistently observed across Patients 119, 209, 210, 214, 215, 221, 223, and 233. In most cases, the F1 score increases rapidly and saturates near its maximum value before approximately 200 ms, indicating stable and early convergence of predictive confidence across subjects (see Fig. 10). A little distinct behavior is observed for Patient 116, where the F1 score begins at a relatively high level (approximately 0.8), subsequently dips during intermediate prediction times, and then recovers toward the end of the sequence, indicating increased temporal variability in predictive stability for this subject (see Fig. 11(a)). An exception is also observed for Patient 228, where the F1 score exhibits a delayed and sharper rise toward the end of the beat, starting from comparatively lower values before rapidly improving at later time points (see Fig. 11(b)).

**Figure 10:**
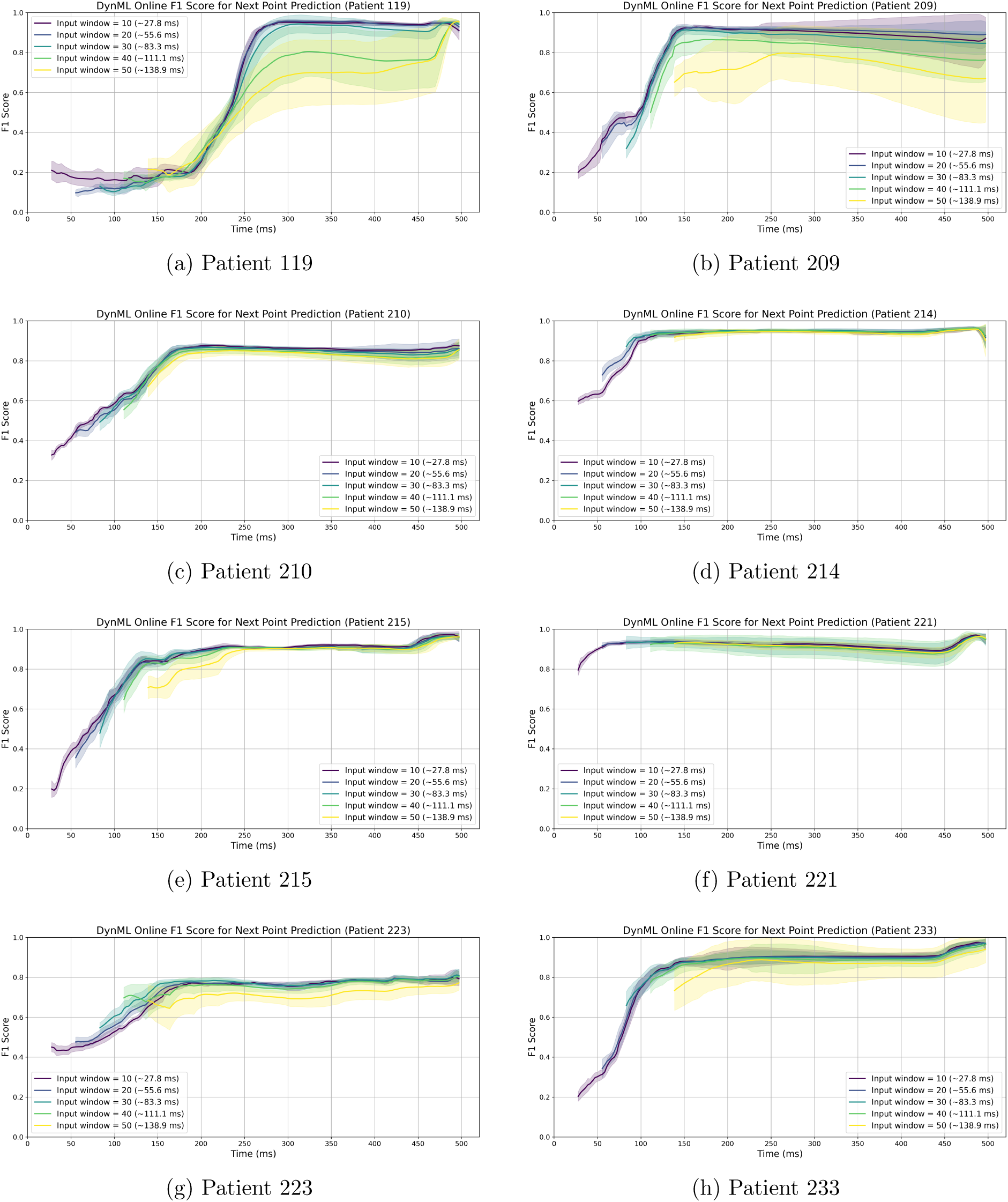
CASCADE online F1 score for next-point ECG prediction across multiple patients under the DynML framework. Mean F1 score trajectories are shown for different input window lengths (10–50 samples) as a function of predicted time (ms) for Patients 119, 209, 210, 214, 215, 221, 223, and 233. Results are averaged over ten independent random seeds. Across patients, the F1 score typically starts at moderate-to-high values and increases with prediction time, showing a rapid improvement phase followed by saturation near 1.0. While slight inter-patient variability is observed in the early prediction phase, all subjects exhibit consistent convergence behavior, highlighting the robustness of the CASCADE framework across heterogeneous cardiac dynamics.

**Figure 11:**
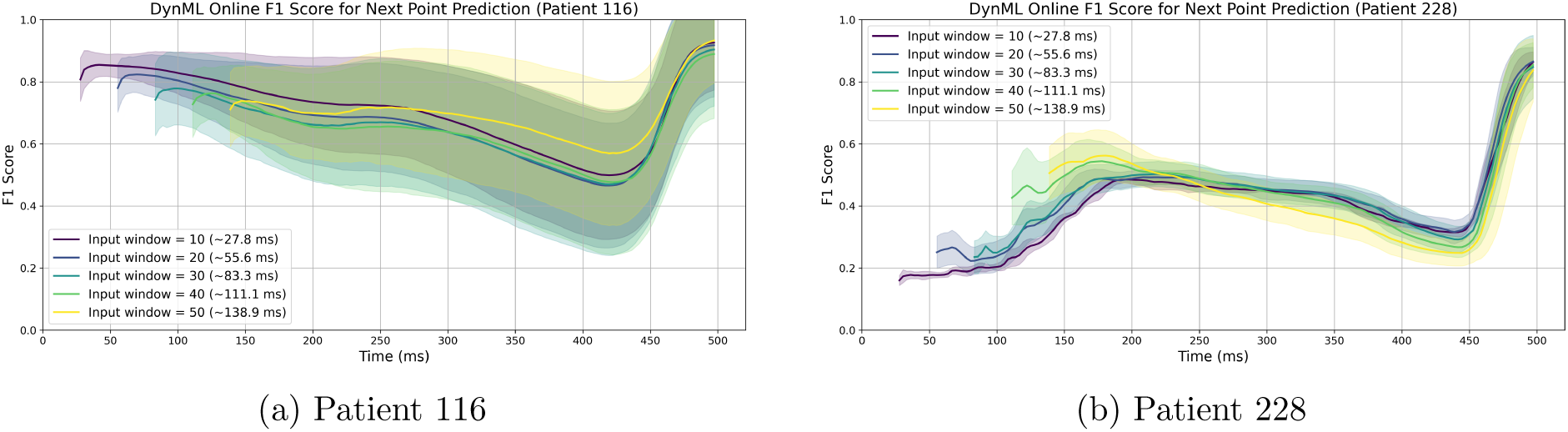
CASCADE online F1 score for next-point ECG prediction in Patients 116 and 228, highlighting atypical inter-patient variability under the DynML framework. Mean F1 score trajectories are shown for different input window lengths (10–50 samples) as a function of predicted time (ms). Results are averaged over ten independent random seeds.

In contrast to the previously discussed patients, the remaining subjects (Patients 118, 200, 201, 203, 207, 208, 213, and 222) exhibit comparatively lower predictive performance, with online F1 scores remaining below approximately 0.6 across most of the prediction horizon. Although a slight increase is observed toward the end of the beat in some cases, the improvement is modest and does not lead to full saturation, indicating weaker predictive confidence and reduced separability in these subjects under the current setting (see Supplementary Fig. 1).

### 3.2 Comparative Online Prediction Performance Across Forecasting Models

The patient-wise analysis presented in the previous section reveals substantial heterogeneity in online predictive performance under the DynML framework. While several subjects exhibit rapid convergence of F1 scores toward high values, others show delayed improvement or consistently lower performance, with F1 scores remaining below approximately 0.6 across most of the prediction horizon. This variability highlights that predictive success is not uniform across patients and suggests that model performance is influenced by underlying differences in cardiac dynamics, signal complexity, and temporal structure.

To systematically evaluate whether these behaviors are specific to the proposed CASCADE (DynML) formulation or are generally observed across standard forecasting architectures, we benchmarked its performance against several widely used neural forecasting models, including a multi-layer perceptron (MLP), a temporal convolutional network (TCN), and a long short-term memory (LSTM) network, using an identical experimental protocol.

All models were trained with the same input window lengths and evaluated in an online anomaly detection setting, where predictive performance was quantified using the F1 score as a function of time following the input window. The selected baselines represent complementary modeling paradigms: the MLP captures static nonlinear mappings, the TCN leverages temporal convolutions to model local dependencies, and the LSTM explicitly models sequential dynamics through recurrent state updates. This architectural diversity enables a systematic comparison of how different learning strategies preserve nonlinear cardiac dynamics in an online forecasting context.

To ensure a fair and comprehensive comparison with conventional forecasting models, we adopted baseline configurations similar to those reported in a recent time-series forecasting study [23], which systematically evaluated MLP, TCN, and LSTM models on physiological time series data. Beyond these reference settings, we conducted an extensive hyperparameter exploration tailored to the ECG anomaly detection task to ensure that baseline performance was not limited by suboptimal tuning. For MLP, TCN, and LSTM models, we systematically varied seven random seeds, batch sizes (64,128,256), and hidden-layer dimensions (100,200,300), while maintaining a fixed training schedule of 100 epochs and a learning rate of 10*^−^*^3^.

We first evaluated all four models for next-point prediction, corresponding to a single-step forecast of approximately 2.78 ms (sampling rate: 360 Hz). For this short prediction horizon, all models exhibited broadly comparable performance across input window lengths, with F1 scores increasing over time as additional post-window observations were incorporated (left panel of Figure 12). The reported trajectories represent the mean F1 score across multiple runs, averaged over different random seeds and hyperparameter configurations. This comprehensive averaging ensures that observed differences between DynML and conventional models reflect fundamental architectural and dynamical properties rather than the choice of specific hyperparameters. Notably, while baseline neural models were sensitive to variations in architectural and optimization parameters, DynML demonstrated comparatively stable performance across all hyperparameter settings. This robustness arises from its reliance on fixed reservoir dynamics and readout-only training, highlighting its suitability for online, patient-specific forecasting and anomaly detection where extensive retraining and hyperparameter tuning may be impractical.

**Figure 12:**
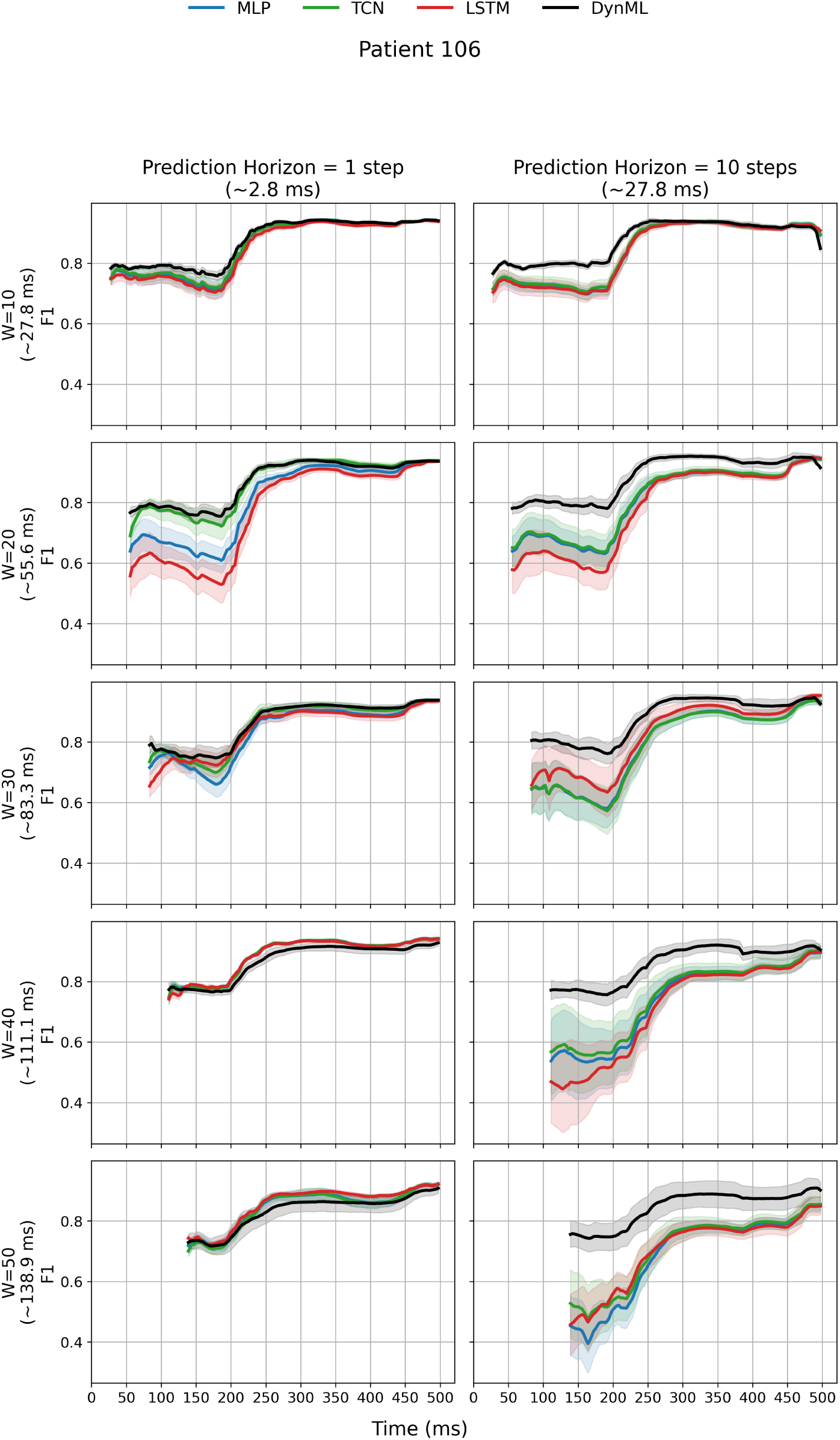
Online F1 score comparison across forecasting models and prediction horizons for a representative subject (Patient 106). Left column: next-point prediction (1 step, ∼2.78 ms), where MLP (feedforward), TCN (temporal convolutional), LSTM (recurrent), and DynML (reservoir-based) exhibit comparable F1 score trajectories across input window lengths. Right column: extended prediction (10 steps, ∼27.8 ms), where the performance of MLP, TCN, and LSTM degrades substantially, while DynML maintains stable and high F1 scores. F1 scores represent the mean ± one standard deviation across multiple runs. The x-axis represents time in milliseconds relative to the end of the input window.

To evaluate robustness under more challenging forecasting conditions, we increased the prediction horizon to 10 output steps, corresponding to approximately 27.8 ms. Under this extended horizon, clear differences emerged: the F1 scores of the MLP, TCN, and LSTM models decreased substantially across all input window lengths, reflecting their reduced ability to sustain accurate online anomaly detection. In contrast, the DynML framework maintained stable and high F1 scores, with trajectories largely unchanged relative to the next-point prediction case (right panel of Figure 12). These results highlight a fundamental distinction between DynML and conventional neural forecasting approaches. While standard models rely primarily on learned input–output mappings that degrade with increasing extrapolation length, DynML employs an entropy-tuned reservoir operating in the chaotic regime. In this configuration, small differences in the input — corresponding to subtle deviations in the cardiac signal — are amplified within the reservoir dynamics, enabling enhanced sensitivity to early-stage anomalies and improved detection robustness.

Importantly, the results shown for Patient 106 (Figure 12) are representative of a broader trend observed across all subjects. As shown in Supplementary Figure 2, similar behaviors are consistently observed for Patients 116, 119, 209, 210, 214, 215, 221, 223, 228, and 233. Across these cases, DynML consistently outperforms MLP, TCN, and LSTM models, maintaining higher and more stable F1 scores under both short- and extended-horizon predictions. While minor inter-patient variability exists in absolute performance levels, the overall pattern remains consistent, reinforcing the robustness and generalizability of the proposed DynML framework for online arrhythmia detection across diverse cardiac dynamics.

In contrast to the previously discussed subjects exhibiting strong or saturating predictive performance, a subset of patients (Patients 118, 200, 201, 203, 207, 208, 213, and 222) demonstrate consistently low predictive performance across all forecasting models. The results for Patient 200 are shown in Fig. 13, while the results for the remaining subjects (Patients 118, 201, 203, 207, 208, 213, and 222) are presented in Supplementary Figure 3. In these cases, online F1 scores remain below approximately 0.6 throughout most of the prediction horizon, with only modest and non-saturating increases observed toward the end of the beat in a few instances. This behavior indicates reduced separability between normal and anomalous dynamics and weaker temporal structure in the underlying ECG signals under the current modeling setup.

**Figure 13:**
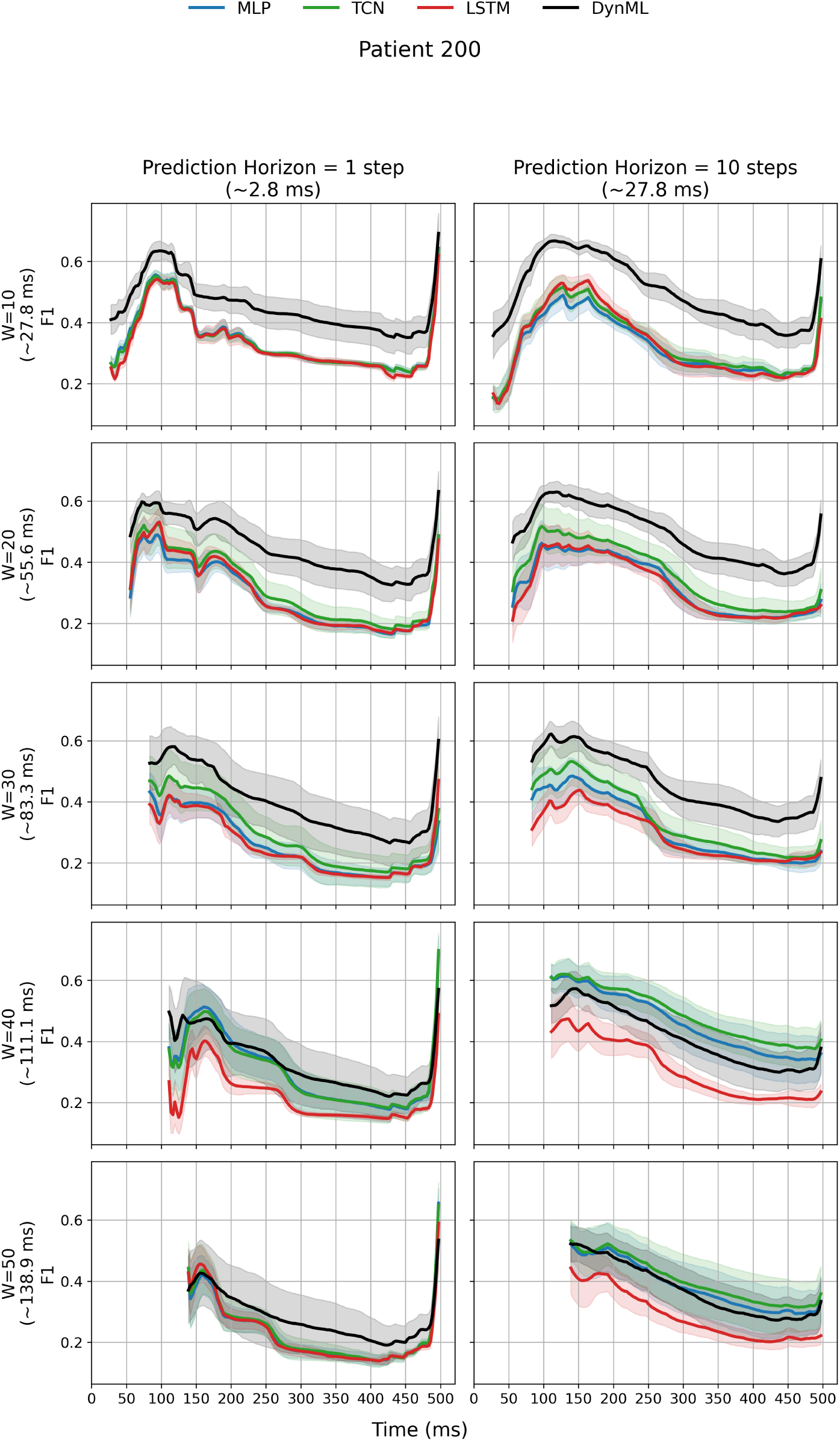
Online F1 score comparison across forecasting models and prediction horizons for a representative subject (Patient 200). Across both next-point (1 step, ∼2.78 ms) and extended (10 steps, ∼27.8 ms) prediction horizons, all models exhibit relatively low F1 scores without strong saturation behavior over time. Despite this overall reduced predictive performance, DynML consistently achieves higher and more stable F1 scores compared to MLP, TCN, and LSTM across input window lengths. F1 scores represent the mean ± one standard deviation across multiple runs. The x-axis represents time in milliseconds relative to the end of the input window.

Despite this overall reduction in absolute performance, DynML consistently outperforms the baseline models (MLP, TCN, and LSTM) across all patients in this group. While all models exhibit degraded predictive confidence relative to the high-performing subjects, DynML maintains comparatively higher and more stable F1 scores across both short- and long-horizon predictions. This suggests that even in regimes characterized by weak signal structure and limited predictability, the reservoir-based dynamics of DynML provide a more robust representation of temporal dependencies, enabling improved anomaly detection performance relative to conventional forecasting architectures.

### 3.3 Early Detection Efficiency Across Forecasting Models

To further quantify the temporal efficiency of each forecasting model, we evaluated the earliest time at which each method reached a range of F1 score thresholds across patients. Specifically, for each subject, we computed the time required for a model to first achieve a given F1 threshold and identified the fastest method per patient at each threshold level. This analysis was performed across multiple thresholds ranging from 0.5 to 0.95 and separately for both short-horizon (1-step) and extended-horizon (10-step) prediction settings.

For the next-point prediction task (1-step, ∼2.78 ms), DynML consistently demonstrated the fastest convergence to clinically relevant performance levels across nearly all subjects. In particular, for lower to moderate thresholds (up to approximately 0.7), DynML was the dominant method, achieving the earliest threshold crossing across all selected patients that reached the respective F1 threshold. This indicates a strong advantage in early detection capability, where rapid attainment of meaningful F1 performance is critical for real-time anomaly identification. At higher thresholds, some variability emerges, with baseline models occasionally reaching the threshold earlier in a subset of patients; however, DynML remains the most frequently selected fastest method overall across threshold levels. Notably, the maximum number of patients for which any baseline method achieved the fastest threshold crossing is limited (e.g., 3*/*13 at an F1 threshold of 0.9 and 1*/*8 at 0.95), while only isolated cases are observed at intermediate thresholds (e.g., 1*/*15, 1*/*15, and 1*/*14 at thresholds of 0.75, 0.8, and 0.85, respectively), further emphasizing the dominant early-time performance of DynML. The results are shown in Fig. 14a. A similar trend is observed under the extended prediction horizon (10 steps, ∼27.8 ms).

**Figure 14:**
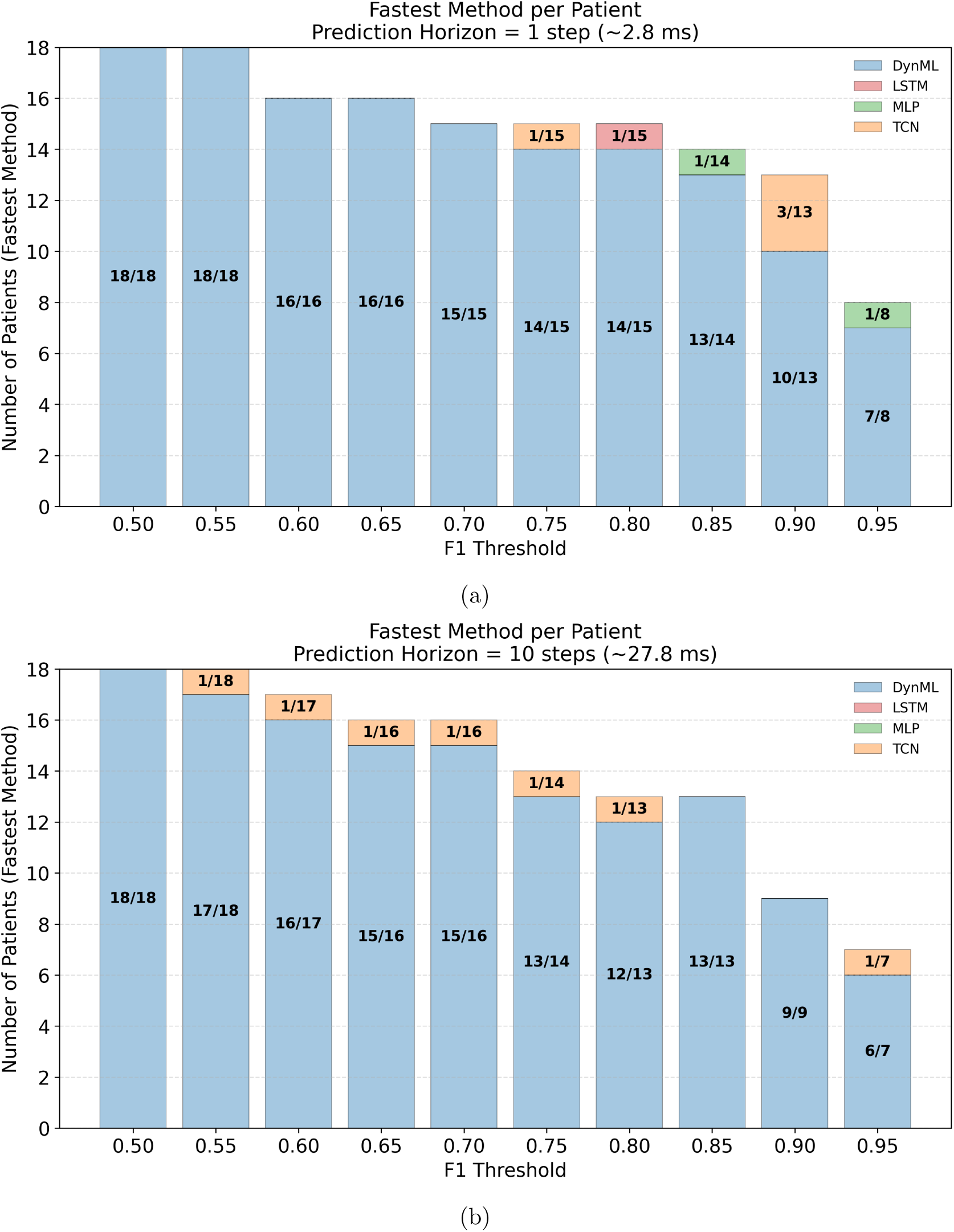
Fastest method comparison based on time-to-threshold F1 performance across patients. (a) Prediction horizon of 1 step (∼2.78 ms) and (b) prediction horizon of 10 steps (∼27.8 ms). The x-axis represents the F1 score threshold, while the y-axis denotes the number of patients for which a given method achieves the earliest (minimum) time to reach that threshold. Each bar is stacked by method (MLP, TCN, LSTM, DynML), indicating the count of patients where that method is the fastest. Numbers inside the bars indicate the fraction of patients (count/total) that reached the corresponding threshold.

While DynML continues to achieve the earliest threshold crossing in the majority of cases, especially at lower and intermediate F1 levels, other models occasionally outperform it in isolated instances. Quantitatively, DynML remains the fastest method for the vast majority of patients across thresholds, achieving 18*/*18, 17*/*18, 16*/*17, 15*/*16, 15*/*16, 13*/*14, 12*/*13, 13*/*13, 9*/*9, and 6*/*7 fastest-threshold cases as the F1 threshold increases. Despite a gradual reduction at higher thresholds, this consistently high dominance highlights the robustness of DynML in rapidly reaching clinically meaningful predictive performance under both short- and long-horizon forecasting settings. The results are shown in Fig. 14b.

Overall, these results demonstrate that DynML not only achieves competitive or superior steady-state F1 performance, but also provides a significant advantage in early-time predictive convergence, which is critical for online arrhythmia detection applications where detection latency is a key factor.

## 4 Discussion

In this work, we introduced CASCADE, an online and personalized framework for cardiac arrhythmia detection that reframes the problem as one of dynamical regime identification rather than static beat classification. By combining entropy-tuned chaotic reservoirs with probabilistic modeling of prediction errors, CASCADE detects arrhythmic events as emergent failures of short-term predictability relative to patient-specific baselines. This formulation directly leverages the nonlinear and intermittently chaotic nature of cardiac electrophysiology and enables robust, real-time anomaly detection without requiring explicit arrhythmia labels during training.

### 4.1 Predictability loss as an early marker of arrhythmic dynamics

A central finding of this study is that arrhythmic events are consistently preceded by a progressive loss of short-term predictability, manifested as statistically significant deviations between predicted and observed ECG dynamics. Rather than relying on morphological pattern recognition, CASCADE detects arrhythmias through cumulative deviations in prediction error likelihoods. This perspective aligns naturally with dynamical-systems theory, in which pathological behavior corresponds to transitions between distinct attractor regimes or to intermittent excursions away from stable oscillatory dynamics [1–3].

The observed sigmoid-like growth of the online F1 score over time reflects the accumulation of dynamical evidence: early prediction errors may be ambiguous, but sustained deviations rapidly push the cumulative log-likelihood below patient-specific thresholds. This temporal integration enables CASCADE to balance early sensitivity with robustness, reducing false positives caused by transient physiological variability. Importantly, this mechanism allows anomaly detection to emerge organically from prediction dynamics, rather than being imposed through ad hoc decision rules.

### 4.2 Role of chaotic sensitivity and topological entropy

Our results demonstrate that the effectiveness of CASCADE is strongly governed by the intrinsic dynamical complexity of the reservoir. Reservoirs operating near critical regimes of topological entropy consistently yielded superior online detection performance, particularly under extended prediction horizons where conventional neural models degrade. This behavior can be understood through the lens of chaotic sensitivity to initial conditions: small deviations in the input signal—corresponding to subtle physiological irregularities—are rapidly amplified by the reservoir dynamics, rendering pathological trajectories linearly separable at the readout level.

This finding provides a principled explanation for why chaotic reservoirs outperform standard recurrent and convolutional architectures in extended-horizon prediction tasks. Whereas deep learning models rely primarily on learned input–output mappings that deteriorate under extrapolation, entropy-rich reservoirs act as dynamical amplifiers whose transient evolution encodes deviations in system behavior [16–18]. Topological entropy thus emerges not only as a descriptive metric of reservoir complexity, but as a practical control parameter for reservoir design. More broadly, this establishes a quantitative link between dynamical systems theory and anomaly detection performance in physiological signals.

### 4.3 Advantages over classification-based arrhythmia detection

Unlike traditional arrhythmia detection methods—which operate as static classifiers trained on labeled datasets—CASCADE requires only normal beats for training and does not depend on explicit arrhythmia annotations. This is a critical advantage in clinical settings, where labeled pathological data are scarce, heterogeneous, and often patient-specific. By learning individualized baselines of normal cardiac dynamics, the framework naturally adapts to inter-patient variability while remaining sensitive to deviations indicative of pathology.

Furthermore, the prediction-based formulation provides interpretability that is largely absent from deep learning classifiers. Prediction errors, likelihoods, and thresholds have clear statistical meaning, and detection decisions can be traced directly to deviations from expected dynamics. This transparency is particularly important for clinical deployment, where trust, explainability, and individualized risk assessment are essential [5, 15].

Cardiac dynamics can vary substantially across individuals, and certain populations—such as athletes—exhibit distinctive heartbeat patterns that differ from the general population. Without a personalized approach, standard anomaly detection methods may misclassify normal variations in these individuals as arrhythmic events, resulting in false positives. Importantly, in this study, CASCADE was evaluated across multiple arrhythmic patterns rather than being restricted to a single type of abnormal beat. This contrasts with many recent approaches that target specific arrhythmia categories; for example, Yuan et al. [24], Gavidia et al. [25], and related studies [26–28] focus exclusively on atrial fibrillation, whereas other methods [12] are restricted to ventricular or supraventricular arrhythmias. By leveraging patient-specific baselines and entropy-tuned reservoir dynamics, CASCADE can robustly detect a broad spectrum of arrhythmic events, making it particularly well suited for heterogeneous populations, including individuals with atypical heart rhythms or multiple comorbidities.

### 4.4 Clinical relevance and broader applicability

The ability of CASCADE to maintain high detection performance under extended prediction horizons suggests its potential utility for early-warning systems in continuous cardiac monitoring. By identifying arrhythmic deviations as soon as predictability begins to fail, the framework may enable earlier intervention compared to methods that rely on fully developed morphological abnormalities. Moreover, because CASCADE operates online and updates its detection statistics in real time, it is well suited for wearable devices and long-term ambulatory monitoring.

Notably, existing wearable and consumer-grade ECG systems, such as smartwatches, have been widely studied for the detection of specific arrhythmias like atrial fibrillation. However, these approaches typically focus on a narrow set of rhythm abnormalities and employ simple thresholding or irregularity detection, rather than providing comprehensive classification of multiple arrhythmia types in a personalized, online manner [29, 30]. In contrast, CASCADE generalizes across diverse arrhythmic events by leveraging patient-specific baselines and chaotic sensitivity, enabling broader applicability in real-time monitoring scenarios.

Beyond cardiac electrophysiology, the principles underlying CASCADE are broadly applicable to other biological systems characterized by nonlinear, multiscale, and intermittently chaotic dynamics. Examples include neural population activity, endocrine regulation, respiratory dynamics, and ecological or epidemiological time series. In these settings, entropy-guided reservoir computing provides a general strategy for detecting regime shifts and anomalies without requiring exhaustive labeled data or detailed mechanistic models.

### 4.5 Limitations

Several limitations of the present study warrant consideration. First, while the Gaussian assumption for prediction error distributions is effective and computationally convenient, real physiological errors may exhibit non-Gaussian tails or time-dependent correlations. Future work could explore more expressive probabilistic models, such as mixture distributions or sequential Bayesian filtering, to further improve robustness. Second, although CASCADE demonstrates strong performance on the MIT–BIH dataset, evaluation on larger and more diverse clinical cohorts—including long-term ambulatory recordings—will be essential to assess generalizability. Additionally, while topological entropy provides a useful global measure of reservoir complexity, it does not capture all aspects of transient dynamics relevant to prediction. Incorporating complementary measures, such as local Lyapunov spectra or information-theoretic transfer metrics, may further refine reservoir design. Finally, integrating CASCADE with physiological models of cardiac electrophysiology could provide deeper mechanistic insight into how specific dynamical instabilities manifest as detectable predictability failures.

### 4.6 Conclusion

In summary, CASCADE establishes a dynamical-systems–based framework for online, personalized arrhythmia detection that leverages chaotic sensitivity and entropy-guided reservoir design. By detecting pathological events as emergent failures of short-term predictability, the framework moves beyond static classification toward a mechanistically grounded view of cardiac anomalies as dynamical regime transitions. This work demonstrates that chaos is not merely a challenge for physiological modeling, but a computational resource that can be systematically exploited for robust anomaly detection in complex biological systems.

## Acknowledgments

This research was supported by the Intramural Research Program of the National Institute of Diabetes and Digestive and Kidney Diseases (NIDDK, ZIA DK075091-13) within the National Institutes of Health (NIH). The contributions of the NIH author(s) are considered Works of the United States Government. The findings and conclusions presented in this paper are those of the author(s) and do not necessarily reflect the views of the NIH or the U.S. Department of Health and Human Services.

## Materials & Correspondence

Correspondence and requests for materials should be addressed to Suvankar Halder.

## Data Availability and Code

All code required to replicate the analyses presented in this study is publicly available at: https://github.com/nihcompmed/CASCADE. The ECG data used in this study were obtained from the MIT-BIH Arrhythmia Database [5], a publicly accessible resource. The dataset can be accessed at: https://physionet.org/content/mitdb/1.0.0/.

