## Supplementary materials for "From Chaos to Care: Personalized AI for Early Cardiac Arrhythmia Warning"

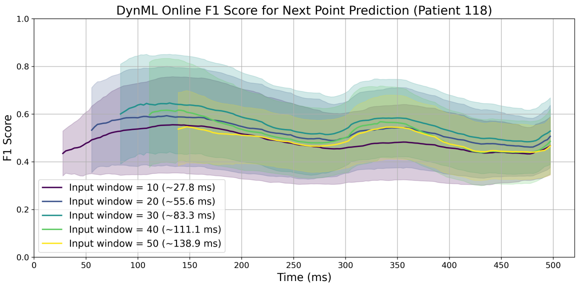


(a) Patient 118

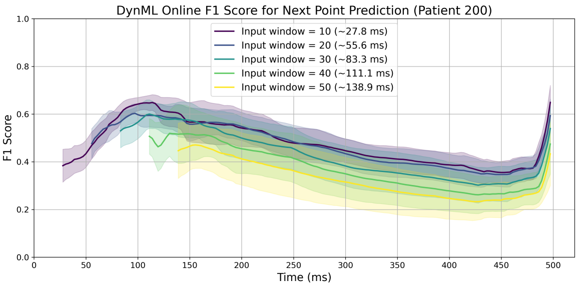


(b) Patient 200

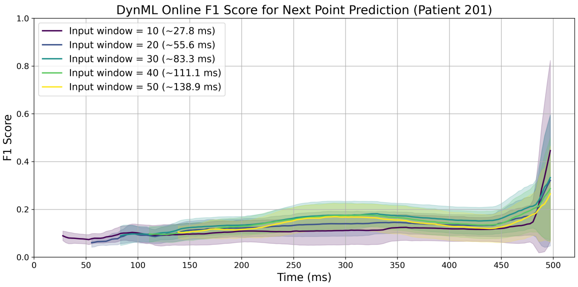


(c) Patient 201

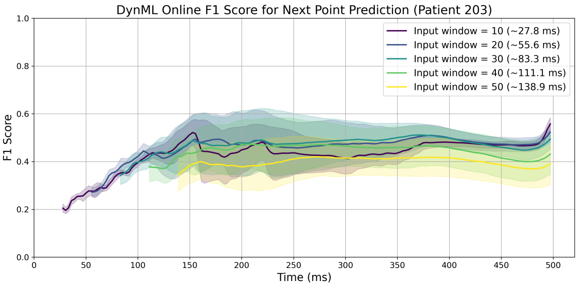


(d) Patient 203

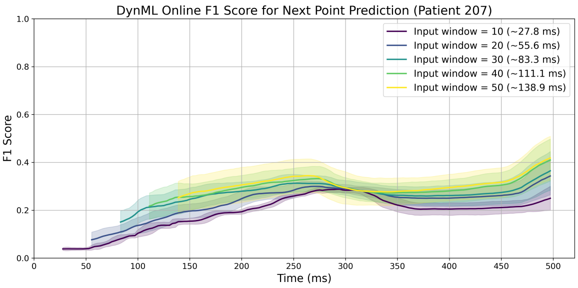


(e) Patient 207

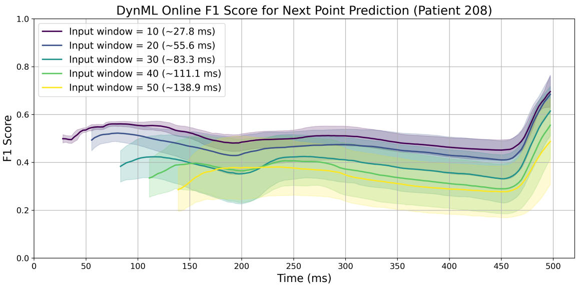


(f) Patient 208

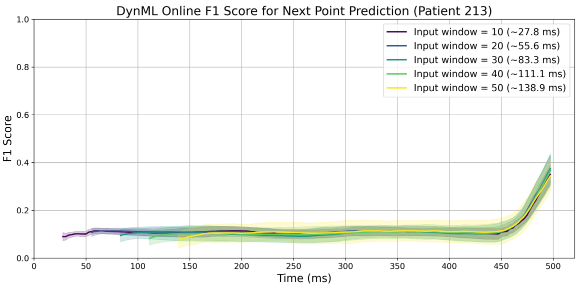


(g) Patient 213

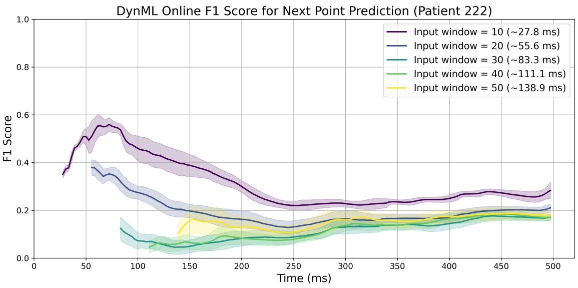


(h) Patient 222

Figure 1: **CASCADE online F1 score for next-point ECG prediction across selected patients under the DynML framework.** Mean F1 score trajectories are shown for different input window lengths (10–50 samples) as a function of predicted time (ms) for Patients 118, 200, 201, 203, 207, 208, 213, and 222. Results are averaged over ten independent random seeds. Across these patients, the F1 score remains relatively low (generally below 0.6) throughout most of the prediction horizon, with only a modest increase observed near the end of the beat in some cases, without clear saturation.

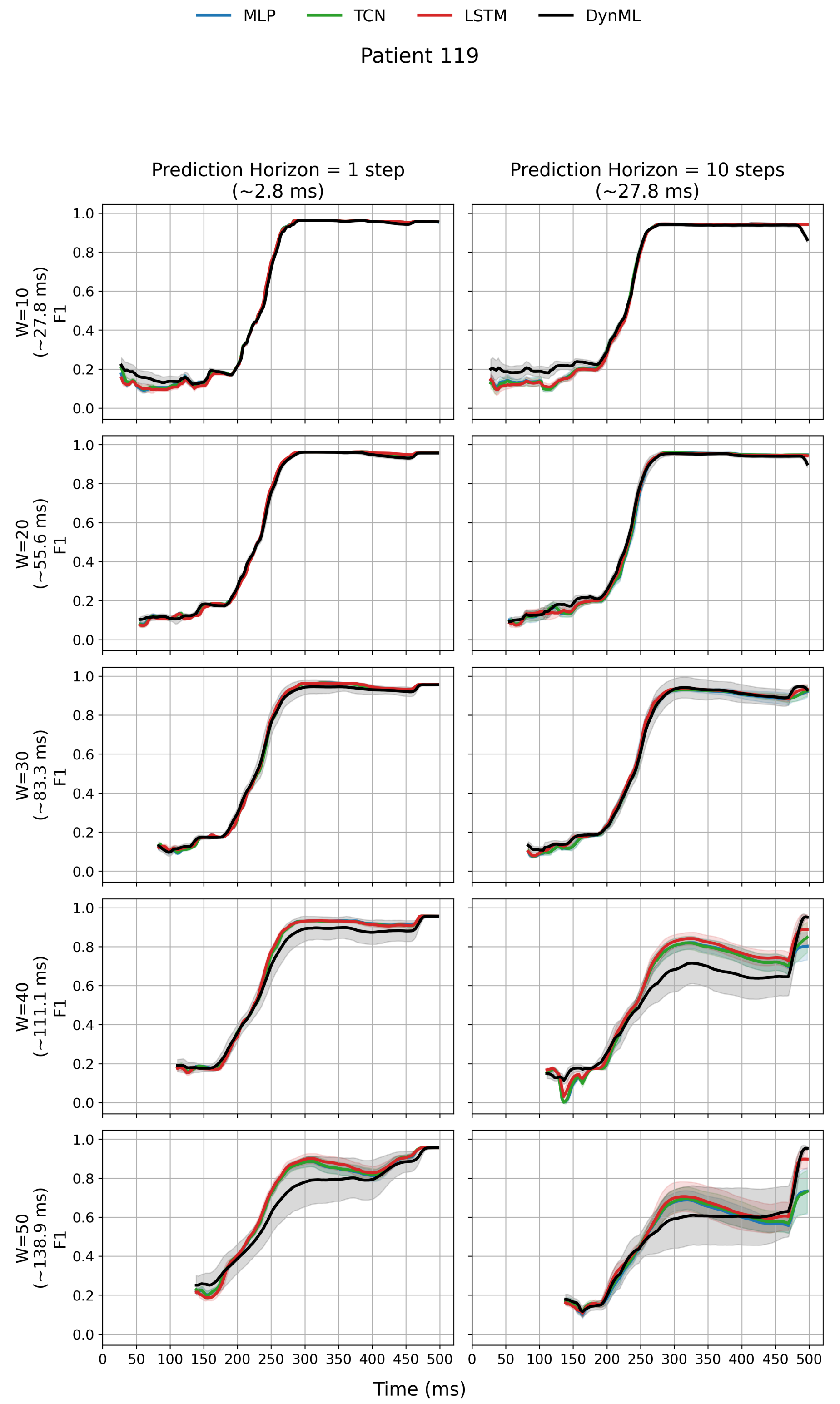


Figure 2: **Online F1 score comparison across forecasting models and prediction horizons for a representative subject (Patient 119).** Left column: next-point prediction (1 step, ∼2.78 ms), where MLP, TCN, LSTM, and DynML exhibit comparable F1 trajectories across input window lengths. Right column: extended prediction (10 steps, ∼27.8 ms), where classical models degrade while DynML maintains stable performance. F1 scores represent mean ± standard deviation across runs.

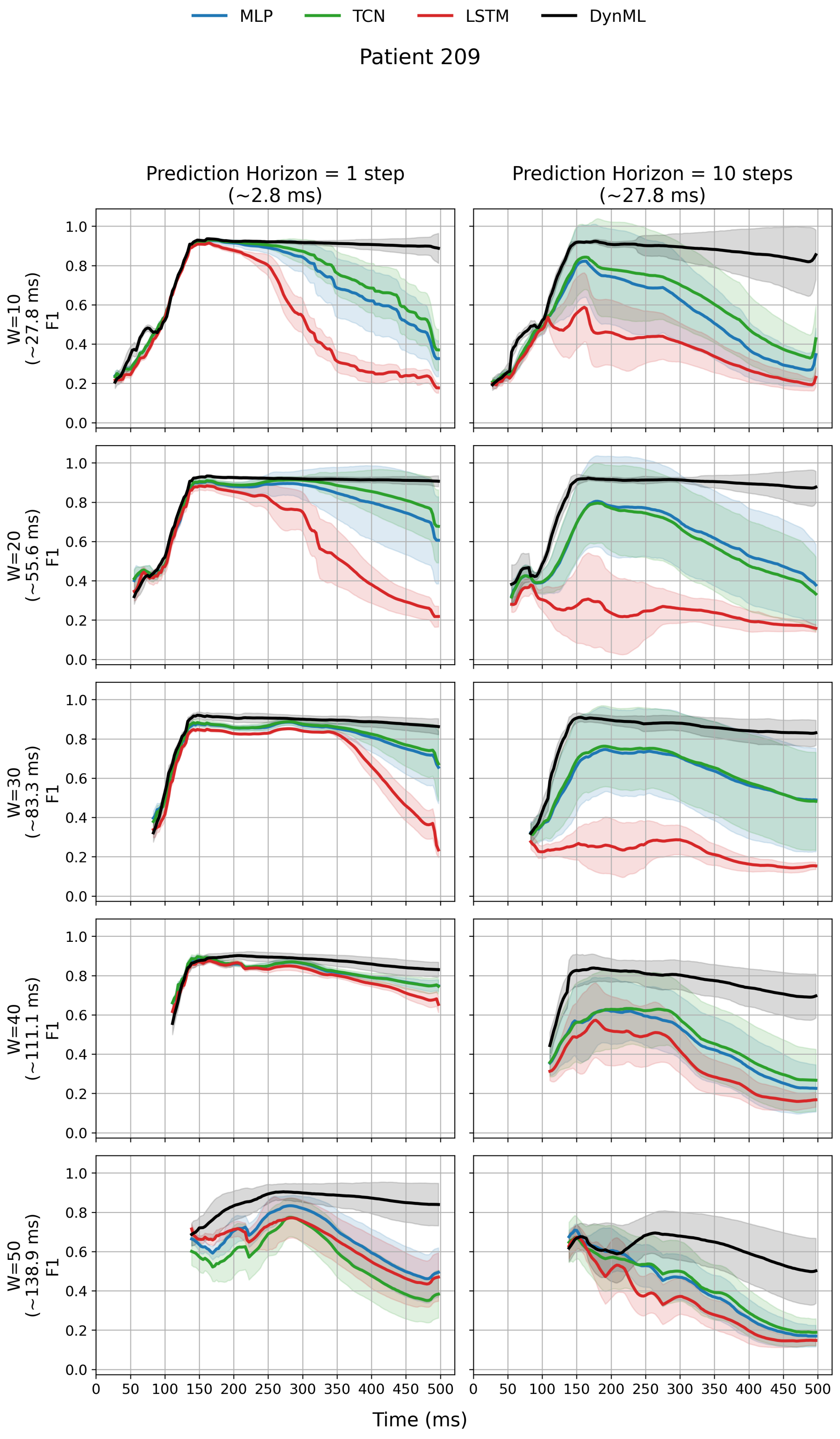


Figure 2: Same description as Patient 119.

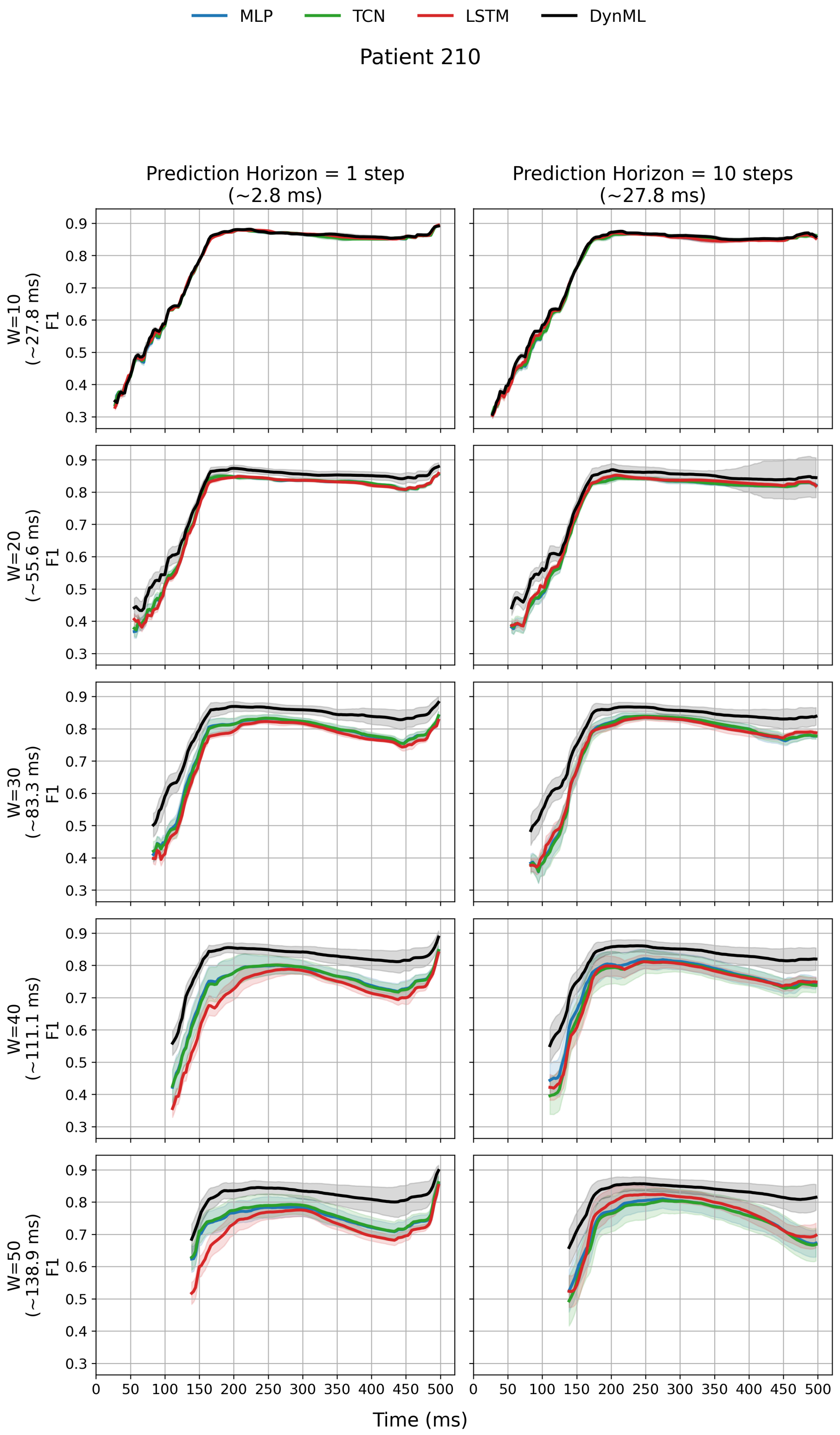


Figure 2: Same description as Patient 119.

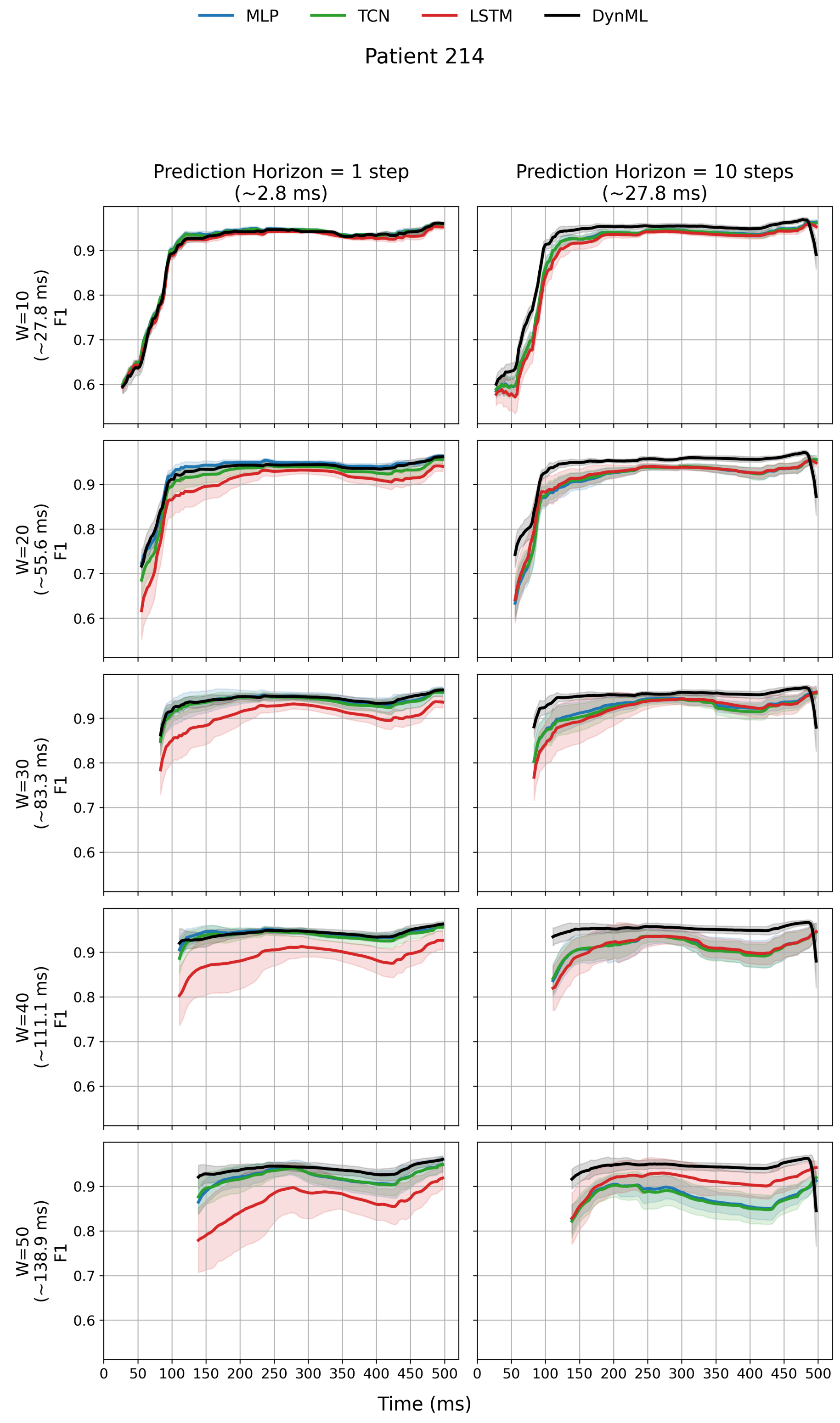


Figure 2: Same description as Patient 119.

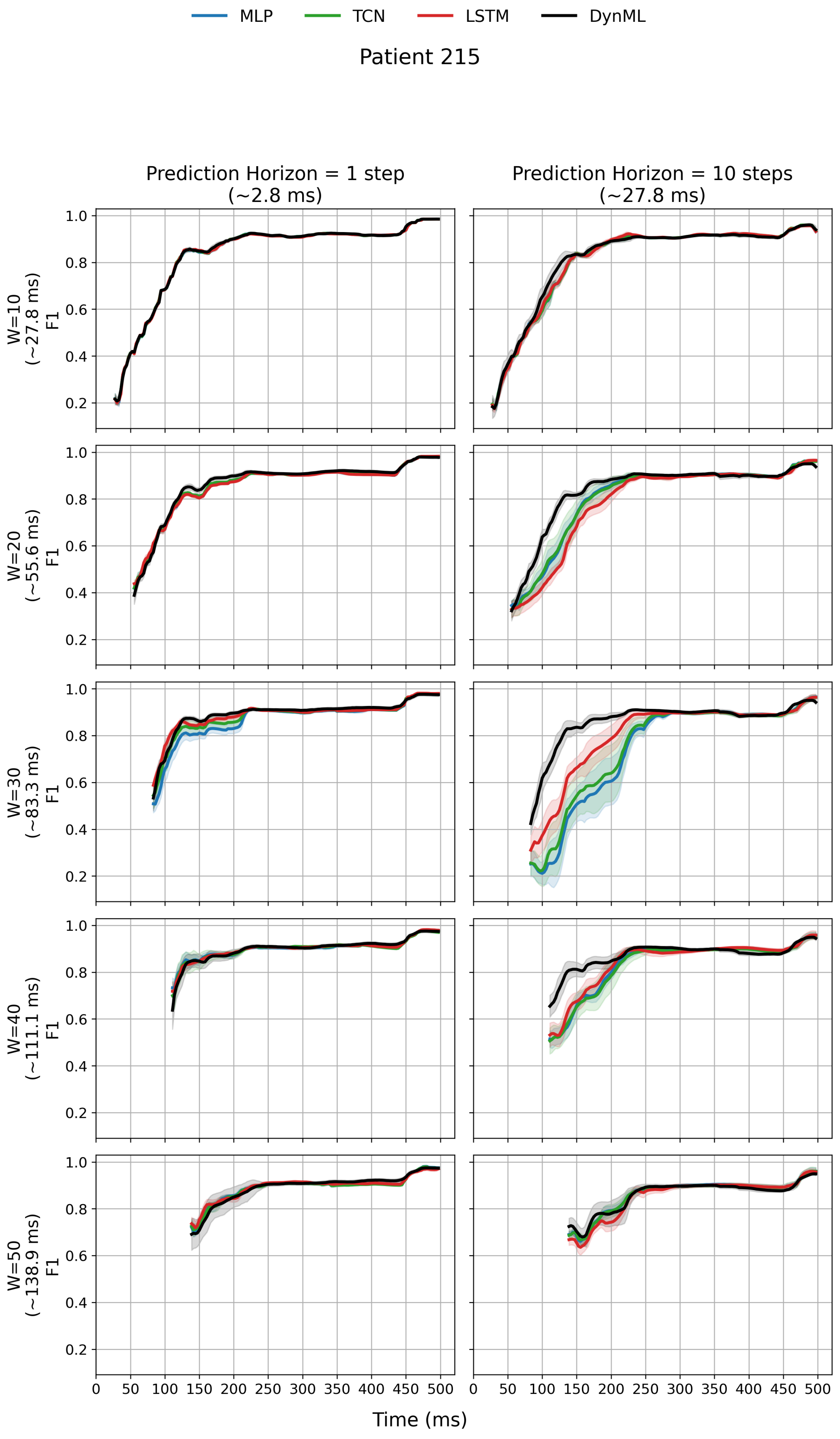


Figure 2: Same description as Patient 119.

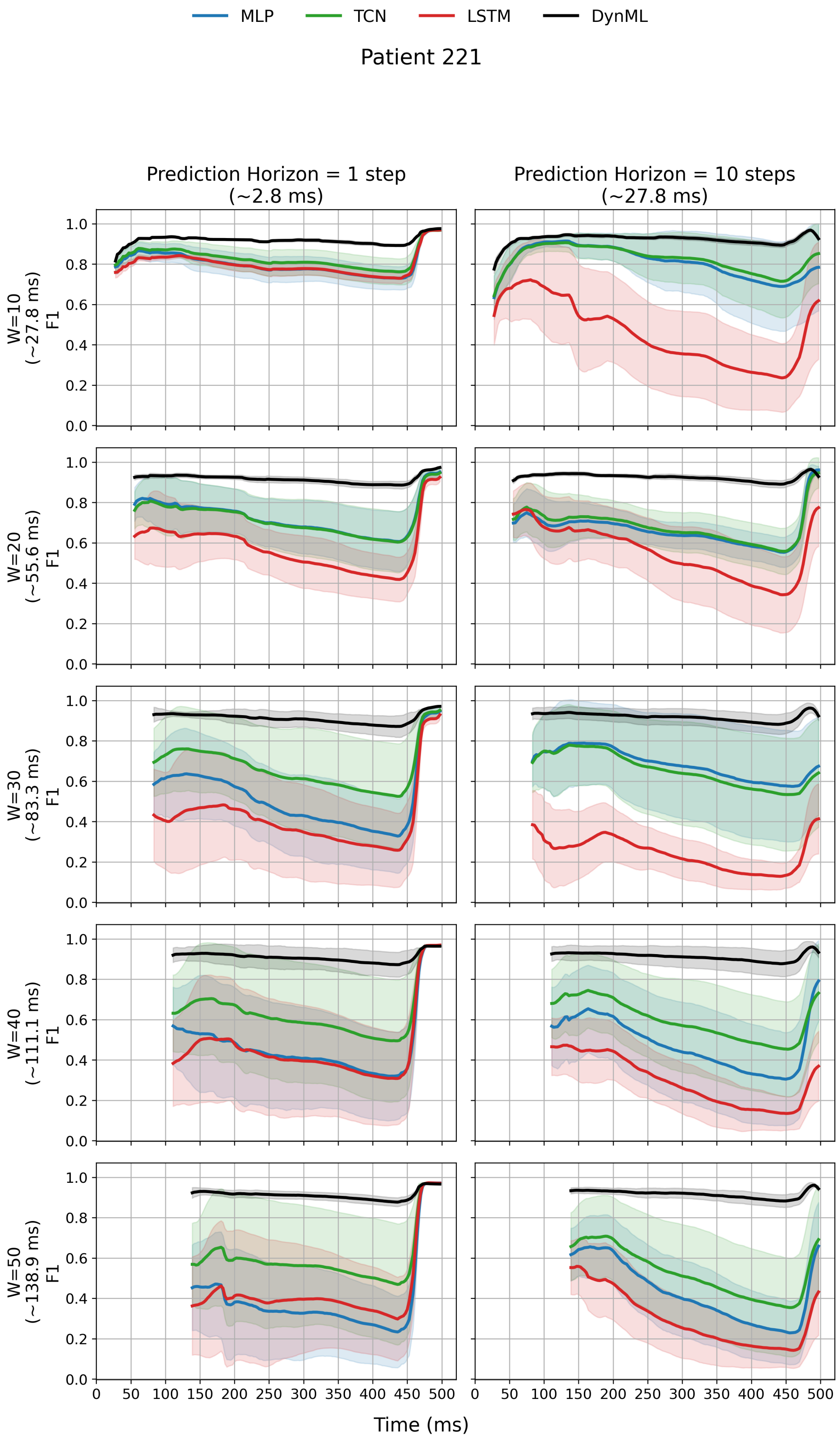


Figure 2: Same description as Patient 119.

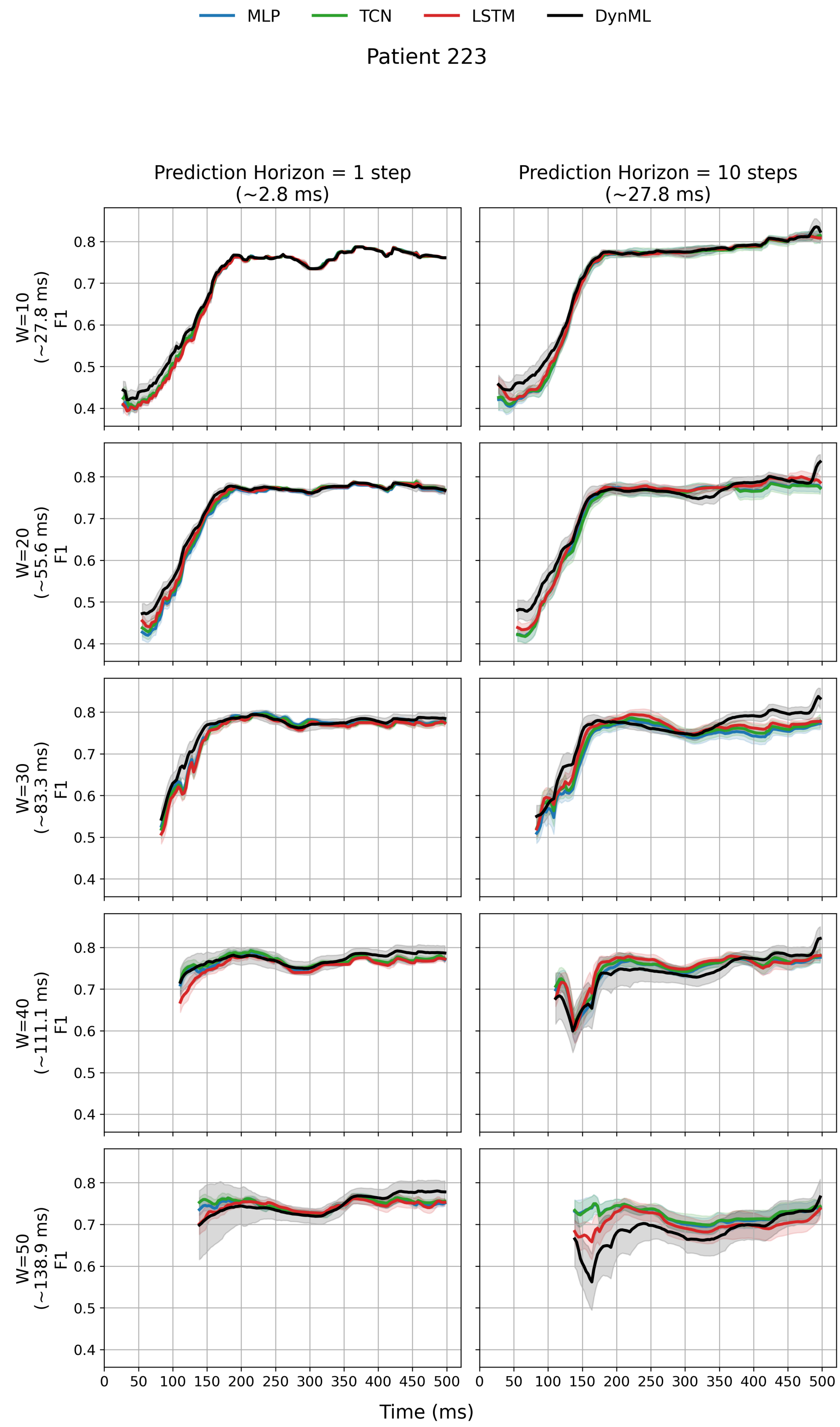


Figure 2: Same description as Patient 119.

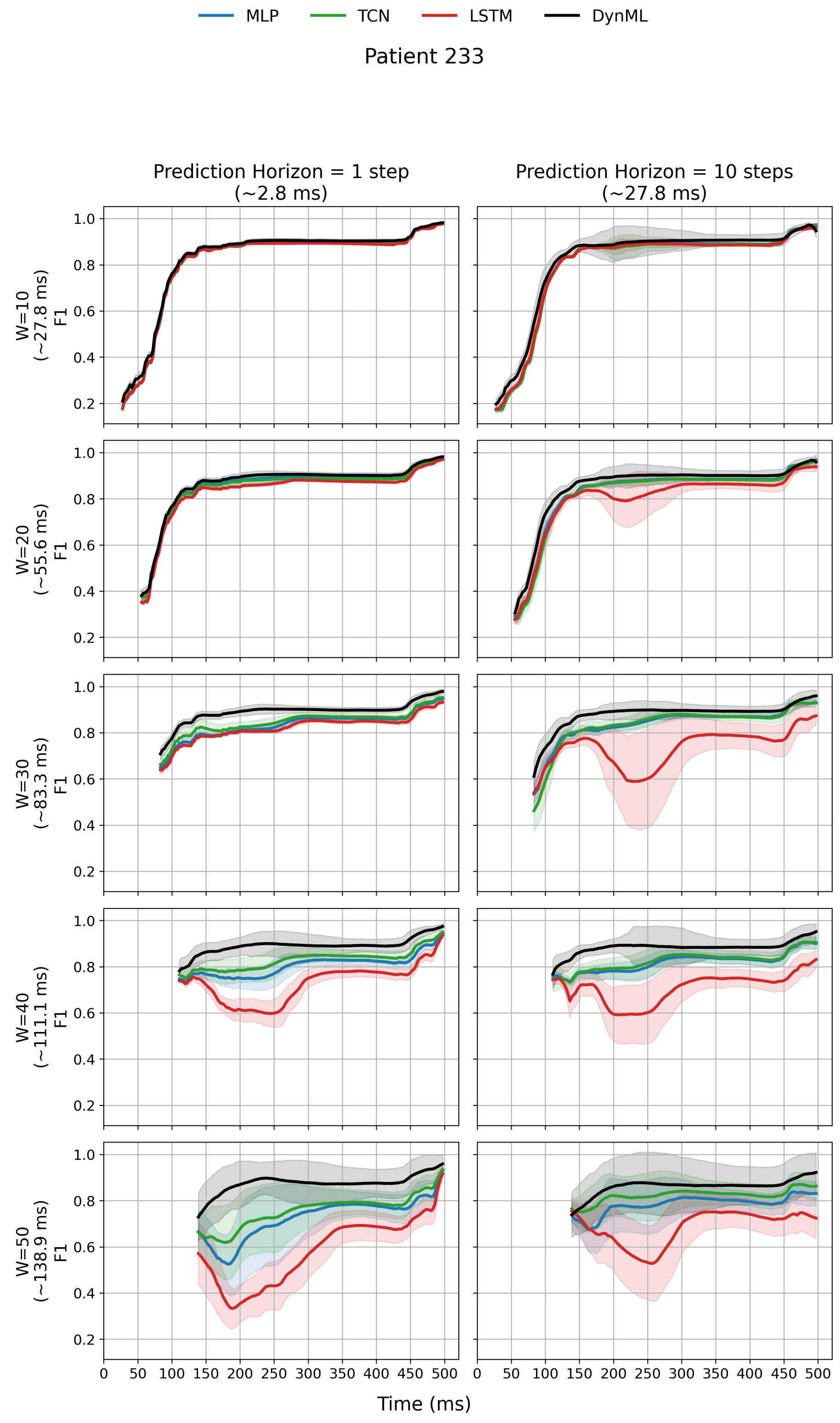


Figure 2: Same description as Patient 119.

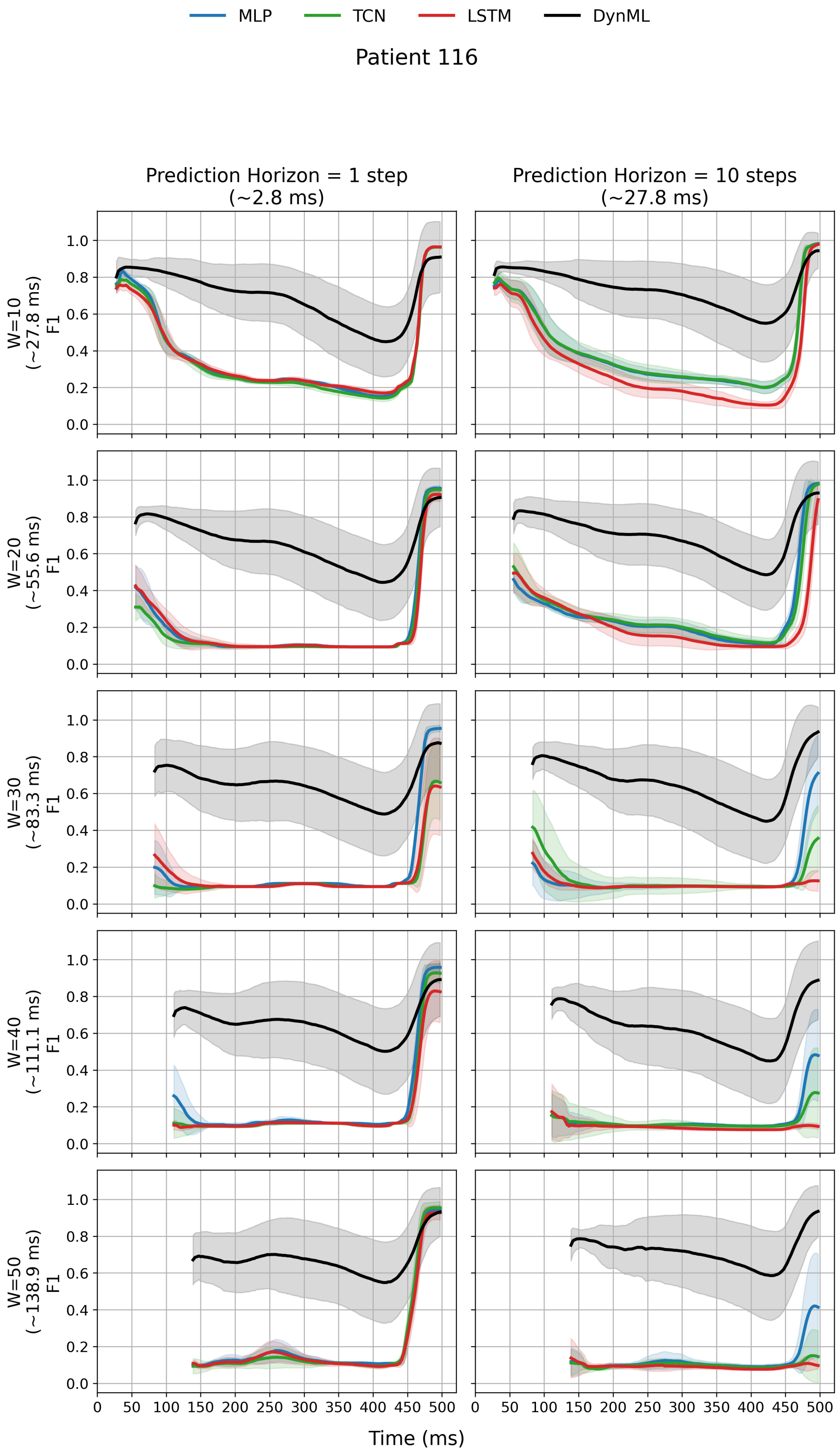


Figure 2: Same description as Patient 119.

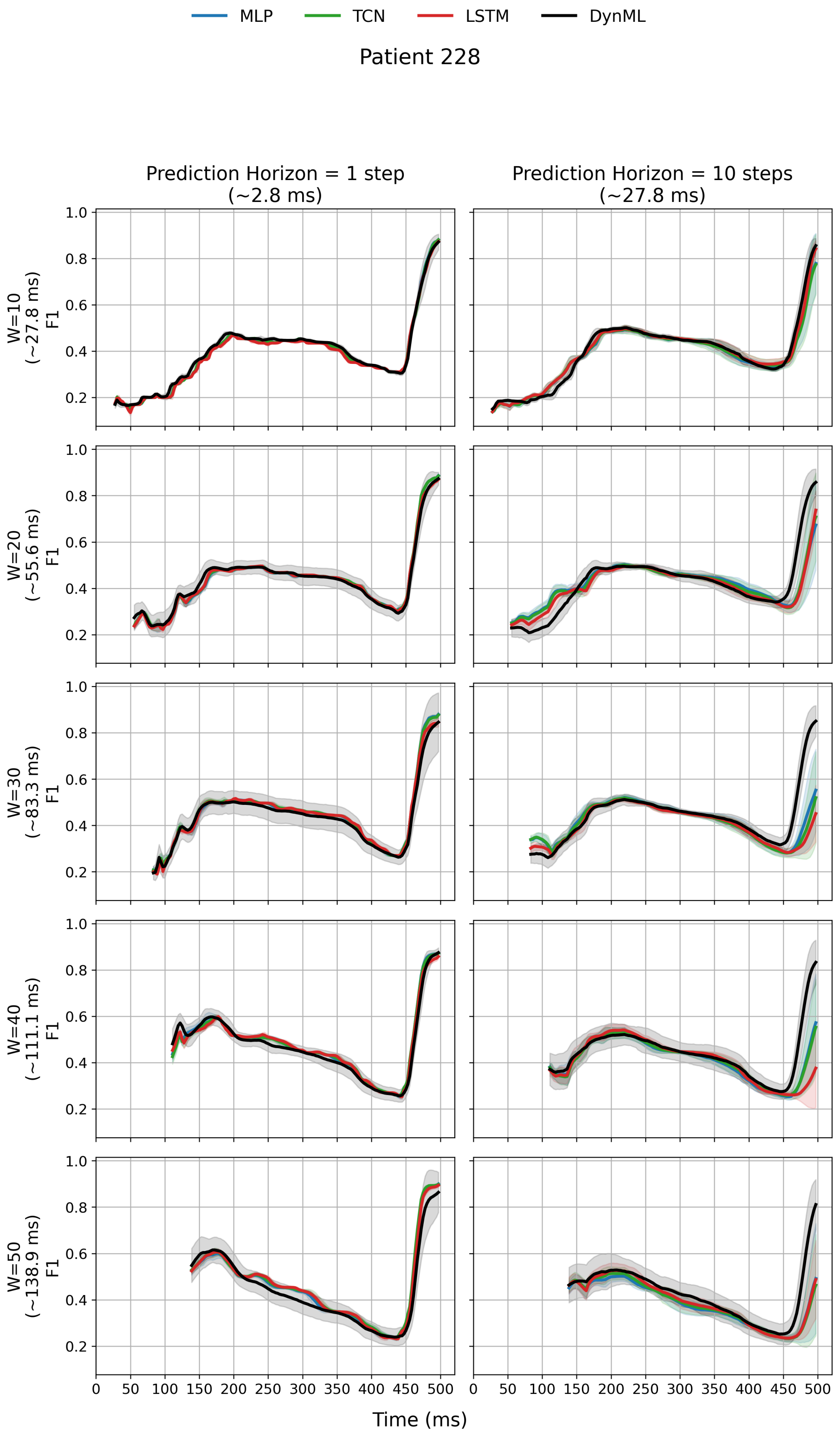


Figure 2: Same description as Patient 119.

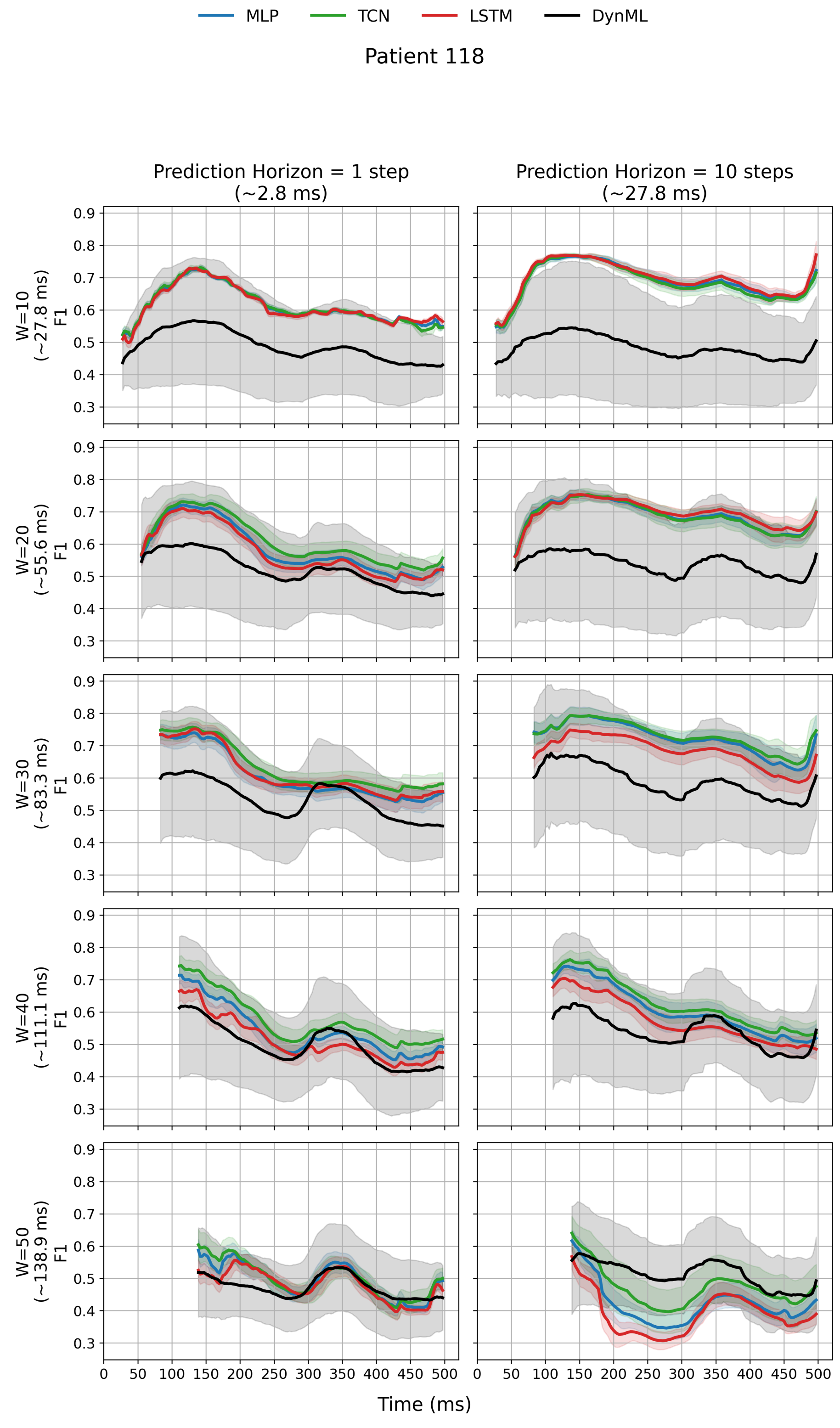


Figure 3: **Online F1 score comparison across forecasting models and prediction horizons for Patient 118.** Left column: next-point prediction (1 step, ∼2.78 ms); right column: extended prediction (10 steps, ∼27.8 ms). MLP, TCN, LSTM, and DynML exhibit comparable F1 score trajectories across input window lengths. F1 scores represent mean ± standard deviation across runs.

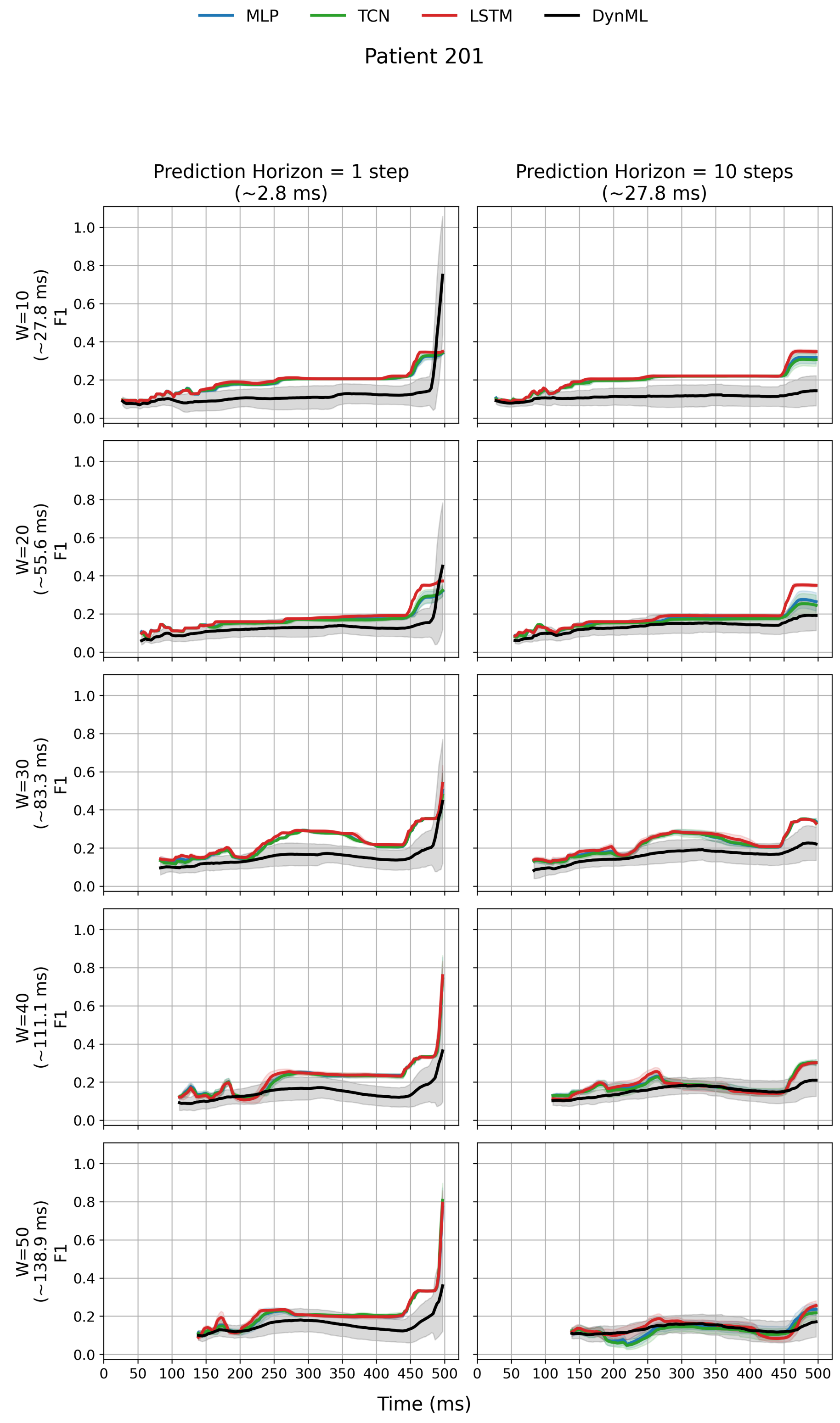


Figure 3: Same description as Patient 118.

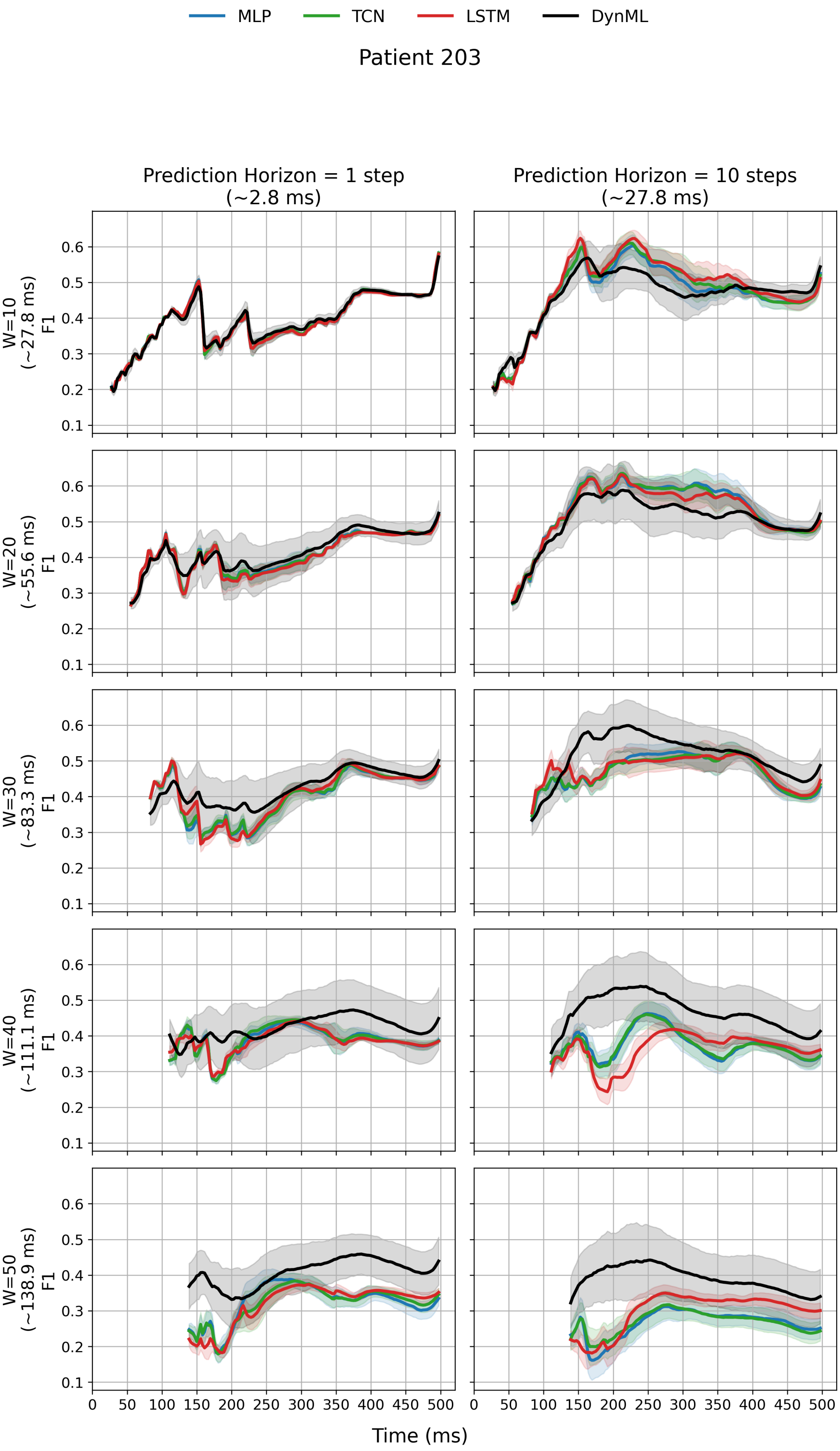


Figure 3: Same description as Patient 118.

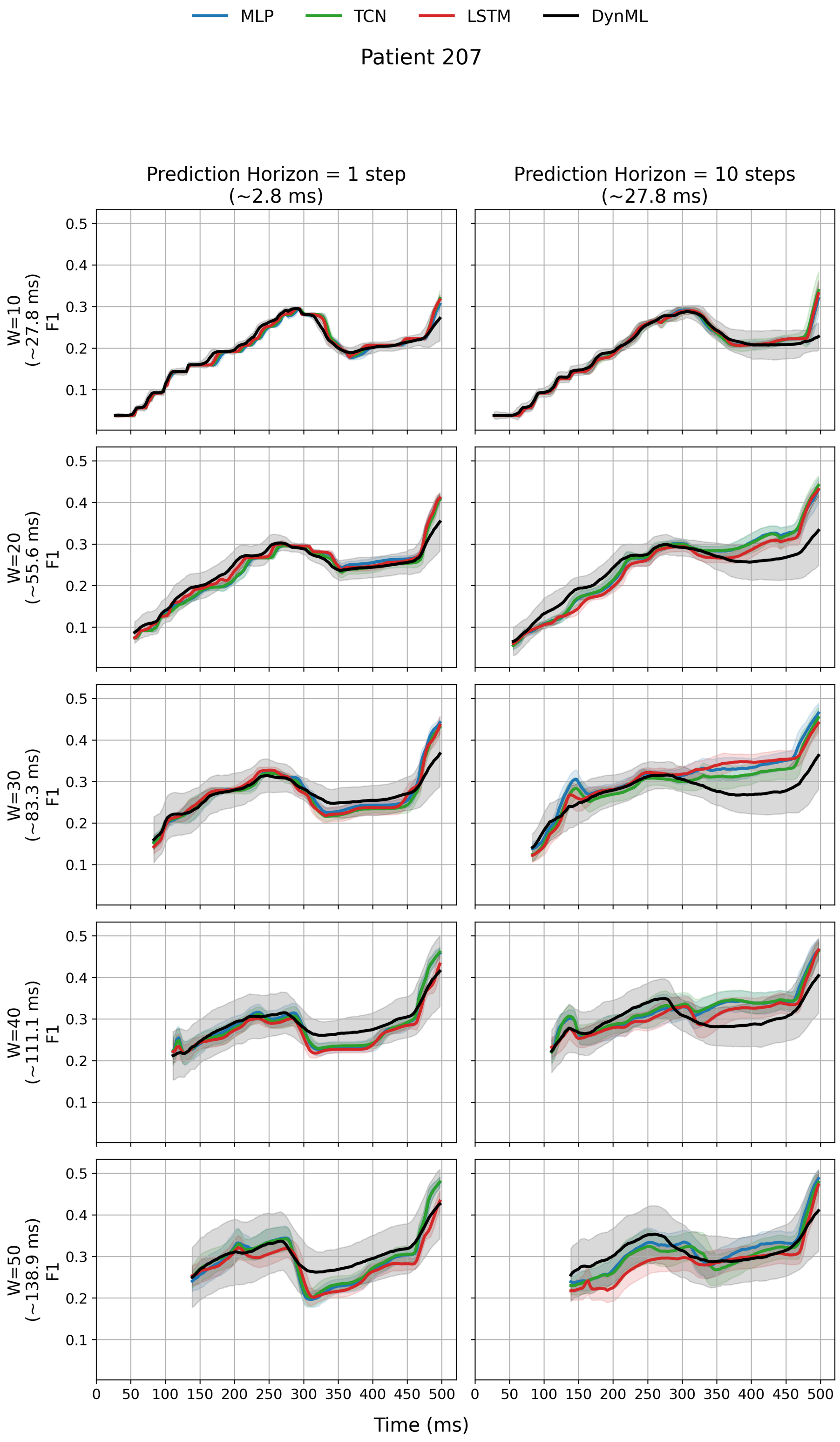


Figure 3: Same description as Patient 118.

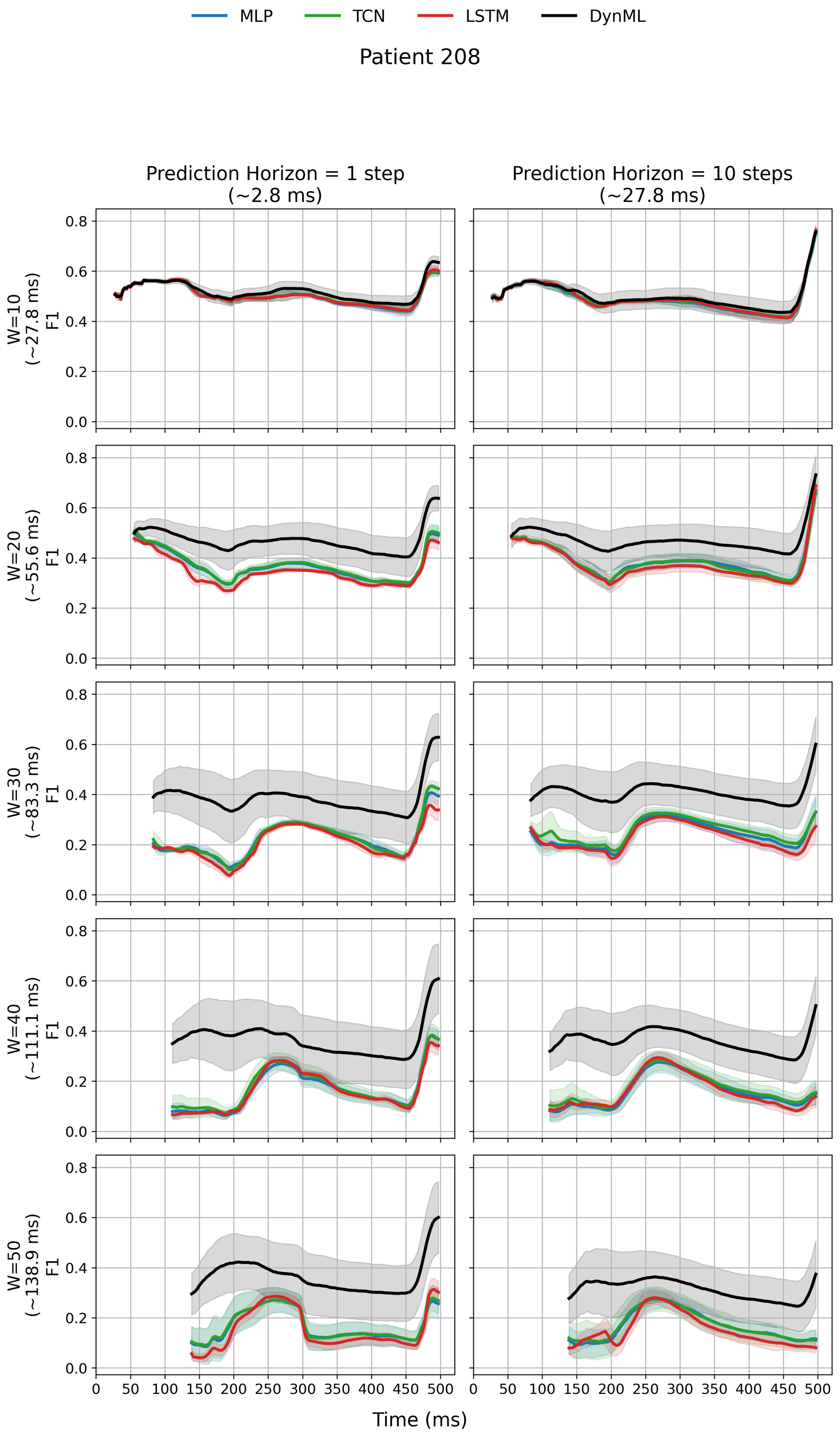


Figure 3: Same description as Patient 118.

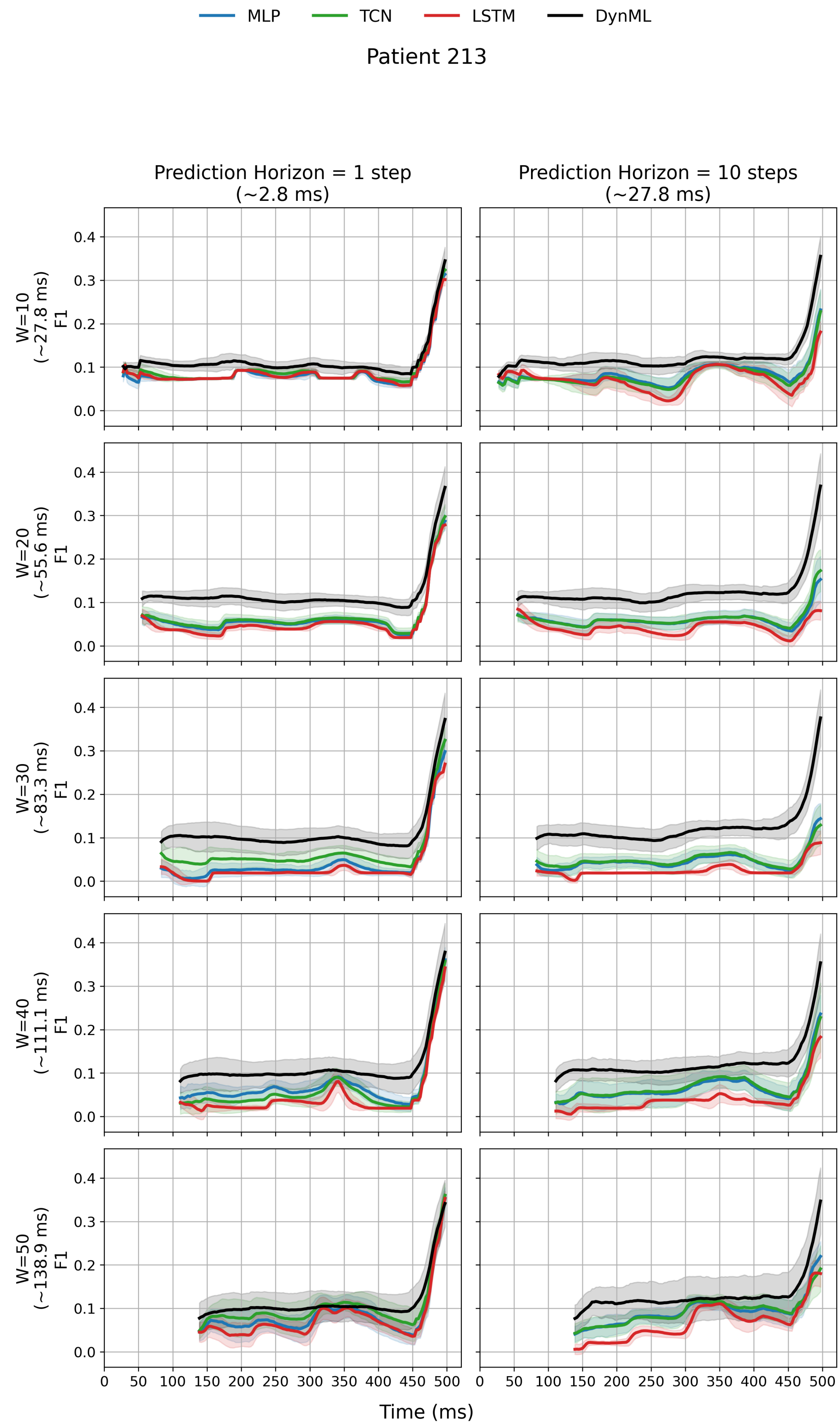


Figure 3: Same description as Patient 118.

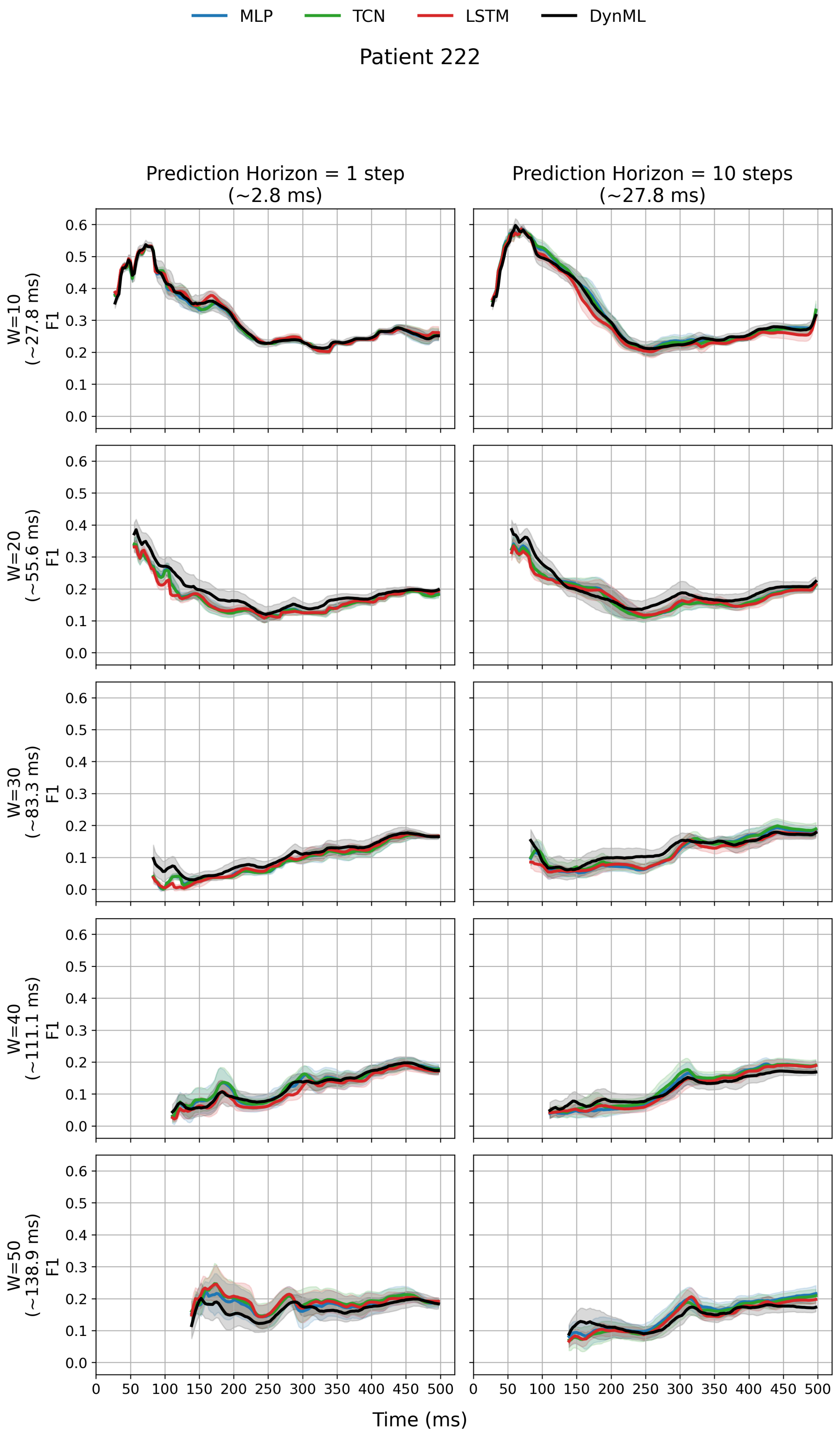


Figure 3: Same description as Patient 118.
